# Immunological and non-immunological features specific to Long-COVID: multiple overlapping aetiologies and potential biomarkers

**DOI:** 10.1101/2025.09.22.25335153

**Authors:** Frederique C Ponchel

**Affiliations:** University of Leeds; Translational research in immune mediate inflammatory diseases, Leeds Institute of Rheumatic and Musculoskeletal Medicine, School of Medicine, The University of Leeds, Leeds, UK; School of Food Science & Nutrition, Faculty of environment, The University of Leeds, Leeds, UK; Rehabilitation, Leeds Institute of Rheumatic and Musculoskeletal Medicine, The University of Leeds, Leeds; Leeds Community Healthcare NHS Long Covid service, Leeds, UK; Department of Immunology, Leiden University Medical Center, Leiden, The Netherlands; School of Medicine, Trinity Translational Medicine Institute, Trinity College Dublin, Dublin, Ireland; Institute of Regenerative Medicine and Biotherapy, INSERM, U1183, University of Montpellier and Clinical Department for osteoarticular diseases, University Hospital Lapeyronie, Montpellier, France; Université Jean Monnet Saint-Etienne, CHU Saint-Etienne, Département de Rhumatologie, and INSERM, SAINBIOSE U1059, Saint-Etienne, France; ELAROS, Electric Works, Sheffield Digital Campus, Sheffield, S1 2BJ, UK; Neuroscience group, Faculty of Biological Sciences The University of Leeds, Leeds, UK; Leeds Institute of Health Sciences, Faculty of Medicine and Health, The University of Leeds, Leeds, UK; Meso Scale Discovery, 1601 Research Boulevard Rockville, MD 20850-3126, USA

**Keywords:** Long-COVID, PRO, Biomarkers

## Abstract

Long-COVID (LC) is a serious clinical condition characterised by debilitating fatigue together with a wide array of symptoms that significantly reduce the quality of life of patients. Currently no holistic or even symptom specific treatment options are available, likely due to both a lack of insight into the disease processes that drive LC symptoms and an extreme heterogeneity in patients’ profiles. We characterised patients and post infection controls, with respect to their immunological profiles with a non-exhaustive panel of biomarkers rationalised based on their potential role in driving symptoms.

We observed that the patients’ symptoms could be grouped into 4 clusters suggesting possible stratification. Systemic inflammation persisted and did not normalize over time in LC. This was not related to persistent SARS-CoV-2 infection, as the presence of circulating N-protein was detected similarly in both patients and controls. No obvious deviation in B-cells and monocytes profiles were observed with minor changes for NK-cells (CD62L+/CD16+/HLA-DR+). Major changes affected CD4+T-cells (and to a lesser extent CD8+T-cells) with respect to exhaustion (PD1+/LAG3+/CD44+), regulation (Treg) and differentiation (naïve/memory-CD62L+). Several candidate biomarkers (cytokines, microRNAs, phosphate metabolism) were present more frequently in LC at high levels and provided information on underlying disease processes. While frequencies of candidate autoantibody+ participants were not different, levels of some antibodies were higher in LC. Yet none of these candidates stood out as a universal biomarker for LC, with the exception of CRP (73% cases), and loss of Treg (50%). However, we confirmed that several overlapping underlying aetiologies may be involved in this complex disease. Specific groups of biomarkers also associated with the 4 cluster of patients. Although to be taken with caution due to small numbers, 3 biomarkers discriminated controls from patients (Treg/CD4+PD1+/CD4+CD161+), others were associated with symptoms recovery (low IL10/IL12/IL4 and high miR766) or deterioration (high CD4+CD38+/ CD8+naiveCD62L+/low IL2) over 12 months. This study provides rational for developing targeted therapeutic strategies as well as biomarkers to stratify LC patients most likely to respond.

## Introduction

Debilitating fatigue and multiple other life-limiting symptoms persisting for >12 weeks and now up to >3 years in many cases, are appearing at an alarming rate of >10% post SARS-COV-2 infections in community-based asymptomatic, and mild/moderate cases and up to ∼30% of severe/hospitalised cases, while less in children (∼1-2%)^[1–3]^. This defines a novel condition publicly known as long-COVID (LC). With >400 million confirmed cases worldwide, >800 million estimated cases^[1–3]^ and given the resurgence of SARS-CoV-2 infections (likely underestimated at >50,000/week by the European Centre for Disease Prevention and Control in 2025), this clearly constitutes a global health emergency^[4, 5]^. As a newly recognized condition, the diagnosis of LC remains challenging. The initial lack of reliable tests to confirm SARS-CoV-2 infection early in the pandemic combined with the heterogeneity of over 70 symptoms^[3, 6, 7]^, led to varying definitions of LC or post-acute sequalae of COVID.

Diagnosis criteria were therefore established in December 2020 in the UK, endorsed worldwide in October 2021. The main symptoms are persistent fatigue for >12 weeks (>80% cases) accompanied by a wide array of other symptoms and a characteristic worsening of these following mental or physical exertion (75% cases)^[6, 8–10]^. LC develops irrespective of the severity of primary infection (including asymptomatic cases), socio-economic background, prior health status, notably affecting previously fully fit, socially active individuals (detailed in supplementary material, self-reported surveys, UK Office for National Statistics and Leeds data Supp-Figure-S2). In addition to significant reductions in quality of life^[11]^, LC has substantial work-related consequences (details in survey), contributing to a significant global economic burden^[12]^.

LC shares symptom similarities to other chronic fatigue conditions with a viral aetiology (such as myalgic encephalomyelitis, ME)^[13, 14]^. Several immune-related aetiologies have been theorised with molecular/cellular mechanisms either as a continuation or as *de-novo* dysfunction of immune-related processes such as immune cell exhaustion due to viral persistence (hidden in various tissue reservoirs), Epstein-Barr virus reactivation, inflammation, autoimmunity and potential epigenetic remodelling of immune cells^[15–19]^. Alternatively, non-immunologic disturbances such as blood clotting and tissue damages, neurological damages, metabolic deficiencies, mitochondrial defects, microbiome dysbiosis^[20–22]^ although in many cases detected in immune cells^[23–26]^. Disentangling cause(s) from consequence(s) remains a major challenge in chronic diseases such as LC. It is therefore crucial to establish whether LC represents a singular mechanistic entity or a combination of symptom clusters with diverse aetiologies, potentially requiring varied treatment approaches. Indeed, most proposed mechanisms are not necessarily mutually exclusive and may overlap or contribute to the range and/or severity of specific symptoms. Alternatively, distinct mechanisms may develop in specific subgroups of patient (e.g. those recovering versus those deteriorating over time), or may occur synergistically, independently, or sequentially. These distinctions will significantly impact the development of pharmaceutical, lifestyle and/or other intervention strategies to reduce the personal and societal burden of this debilitating condition.

To investigate the multifaceted processes potentially driving LC, we recruited LC patients (LCP) to compare with fully recovered post-COVID controls. We selected a panel of molecular and cellular features associated with various possible mechanisms (viral persistence, T-cell exhaustion, differentiation, inflammation, autoantibodies, fatigue biomarkers and microRNAs). Our primary aim was to identify features specific for LC, distinguishing them from those present or returning to baseline in controls, and secondly, to correlate these features with symptom profiles. Our ultimate objective was to provide a list of potential biomarkers associated with patients’ symptoms and trajectories.

## Methods

### Extended methods are provided in supplementary material

#### Patient/control recruitment

One group (n=93) was recruited specifically for biomarker investigations in Leeds (REC-21/EM0112) via the dedicated Leeds long-COVID service/pathway set up in NHS-England. Inclusion criteria were age 18-65 and persistent fatigue for >12 weeks. Prior SARS-CoV-2 infection was verbally verified. A Fatigue Impact Scale (FIS) was collected at the time of blood sampling. Demographic/symptom questionnaire (PCR proof of infection if available) were retrieved from a LC specific questionnaire developed in 2020 (C19-YRS-PRO/PROM (https://c19-yrs.com/the-c19-yrs/) via the NHS-LC pathway. A second group (n=20) was recruited in Saint-Etienne from a regular chronic fatigue clinic (ID NCT05899595). University Staff who had fully recovered from COVID-19 without experiencing fatigue were included as a no-fatigue controls (NFC, n=41). All participant signed an informed consent form.

#### Flow cytometry

was performed on cryopreserved cells. For cell subsets known to be correlated with age (naïve, memory, regulatory) data were normalised (Supplementary figure-S2). NLRP3+ASC flow cytometry methods were previously described^[27]^.

#### Cytokine/CRP quantification

was performed using high-sensitivity MSD chemio-luminescence assays.

#### N-Protein

was detected using a MSD single-plex assay in serum.

#### Autoantibodies

RF-IgM, anti-citrullinated peptide antibody (ACPA), Anti-CarP antibodies were detected using ELISA and antinuclear antibodies (ANA) using immunofluorescence.

#### microRNA RT-qPCR

Total RNA was extracted from 200 µL of serum and converted into cDNAs. Levels of miR expression was determined using hsa-mir-766, hsa-mir-144, hsa-mir-21, hsa-mir-671 off-the-shelf assays

#### Biomarkers of the phosphate metabolism

detailed in supplementary methods.

#### Statistical analysis

Data exploration used univariate analysis (Mann-Whitney-U, Chi^2^). To assess collinearity between variables we used the Cluster-3 algorithm (Stanford University 1998-99) based on Spearman correlations on Log-transformed data. Results are displayed using heat maps of biomarker levels (TreeView). This algorithm is not relaying on statistical significance. Correction for multiple testing was not applied, and p-values (<0.05) were considered trends in most analysis. SPSS V29 and GraphPad Prism V9 were used. Logistic regressions to discriminate LC from NFC, is described in supplementary material.

## Results

### Patients and no-fatigue controls post-infection

The cohort of LCP (LCP, n=93 UK and n=20 France) and no-fatigue controls (NFC, n=41) were matched as far as possible for age and sex (Figure-1A). The symptom duration since initial COVID-19 infection was comparable between groups. The severity of initial infection was similar. The Fatigue Impact Scale (FIS) scores were markedly elevated in LC. Importantly, FIS decreased with age in NFC (r=-0.333, p=0.030, n=41) but not in LC (r=+0.080, p=0.520, n=65).

**Figure 1:**
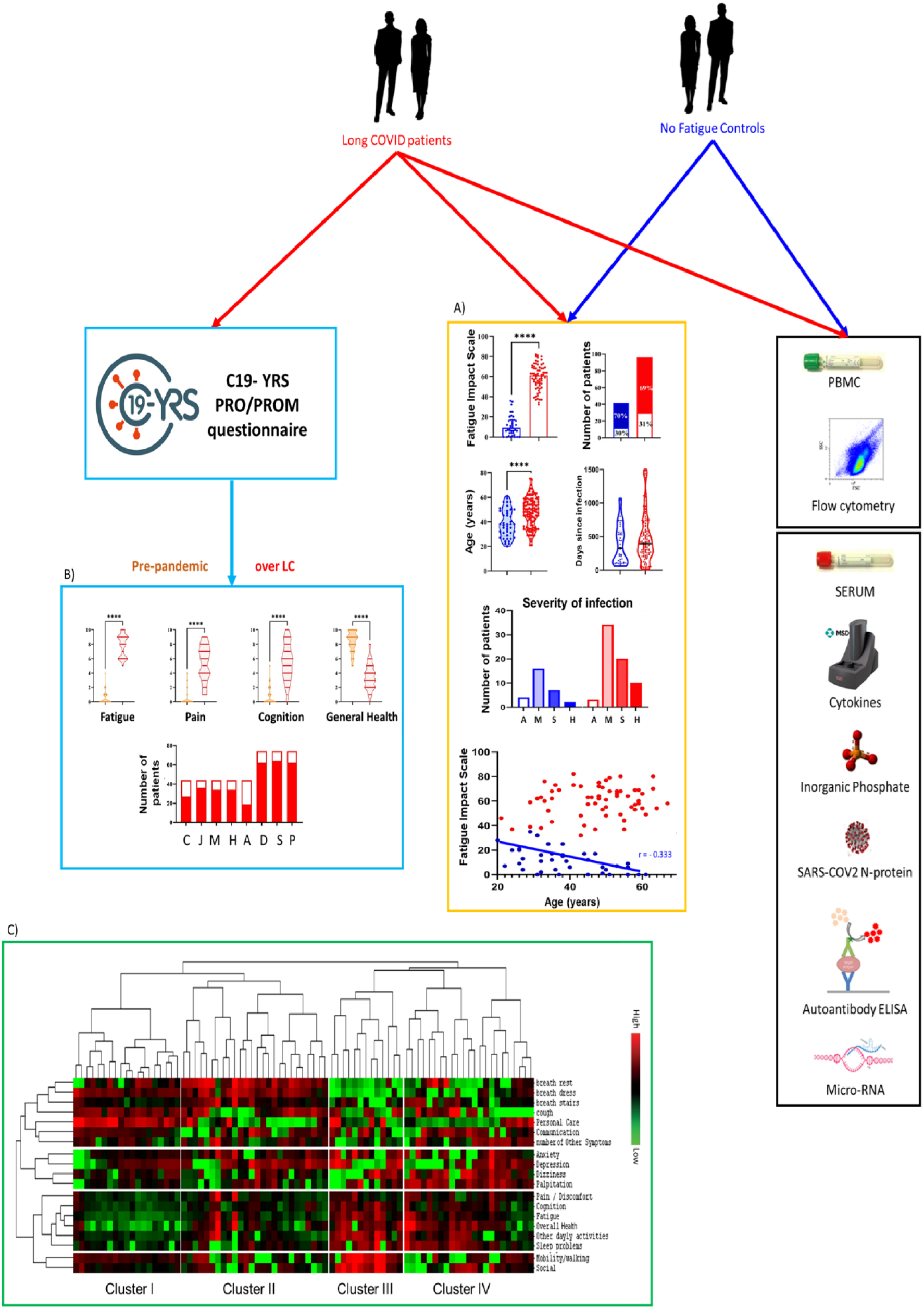
Project overview. Long-COVID patients (LC) and no fatigue controls (NFC) were included in this study. 93 LC (red) and 41 NFC (blue) donated blood (55 mL for PBMC/Serum for 63 LC/41 NFC and 9 mL serum only for 26 LC which were used in various biomarker panel/platforms (flow cytometry, MSD-chemiluminescence platform, biochemistry/ELISA assay for various markers, and qPCR for miRNAs). A) Described demographics. Male are presented as open histograms and female as closed. Severity of the primary infection is classified as asymptomatic (A), mild/moderate 2-5 days felling sick (M), severe >2 weeks sick (S) and hospitalised cases (H). **** p<0.0001 by MWU. Of note, the younger the NFC, the higher was their perception of fatigue (fatigue impact scale FIS) despite considering themselves healthy. Spearman correlation coefficient rho displayed on the graph. B) Data from the C19-YRS-PRO/PROM questionnaire were retrieved from the Long-COVID NHS-pathway and used when withing 30 days of the blood samples (n=82). Scores for fatigue, pain and cognition (0=>10 scale) and general health (10 => 0 scale) are plotted pre-pandemic (orange) and over LC (red). Location of pain was available in 42 cases: chest (C), joint (J), muscle (M), headaches (H) and abdominal (A). Dizziness (D), sleep disturbances (S) and palpitation (P) were also recorded (n=72). **** p<0.0001 by MWU. C) Symptom patterns were analysed using the Clsuter-3 algorithm (unsupervised clustering approach based on Spearman correlations of log-transformed symptom severity scores, not relying of significance) visualizing results on a heatmap of the scores (log transformed). Symptoms segregated into 3 groups and patients into 4 clusters (I to IV).

We analysed data from the C19-YRS-PRO/PROM questionnaire in 82 LCP (not available in NFC), when captured within 30 days of the blood samples collection. Symptom profiles were consistent with those expected for LC (Figure-1B, and Supp-Figure-S3) all reporting fatigue scores >5 (0-10 scale). Patients rated their overall health as high pre-pandemic (median=8, range 5-10) and poor during LC (median=3, 0-6). Despite the relatively small sample size, our cohort is highly representative of the broader LC population.

### Symptom profiles

Considering the heterogeneity of PRO/PROM reported by LCP, grouping them in prototypic profiles is paramount to understand their potential common origin and/or mechanism. Cluster-3 algorithm was used to cluster symptoms into 4 main areas (unsupervised) (Figure-1C):

- 1^st^ group reflected overall respiratory function (at rest/dressing/climbing stairs/cough) with personal care/communication
- 2^nd^ group included anxiety/depression/dizziness/palpitations
- 3^rd^ group encompassed the main symptoms (fatigue/pain/cognition/sleep/overall-health/impaired daily activities)
- 4^th^ group associated mobility with social life issues.

Patients segregated into 4 distinct clusters.

`o` Cluster-I exhibited high respiratory issues but relatively low fatigue/pain.
`o` Cluster-II had severe symptom group-1 only
`o` Cluster-III displayed highly impacted symptoms in group-3/4 and markedly lower severity in group-1/2.
`o` Cluster-IV had severe group-3 symptoms.

We then proceeded to analyse a large number of serum biomarkers and cell subset individual and/or per platform and then proceeded to multi-marker analysis and associations with patient data.

### Systemic inflammation and circulating cytokines

Ample evidence link inflammation with chronic fatigue^[28, 29]^. Serum levels of CRP and cytokines associated with pro-inflammation (IL-1-beta/IL-6/IL-10/IL-17A/TNF), immune polarisation (IL-2/IL-4/IL-12p70/IL-13/IFN-γ), and pain (IL-8)^[30, 31]^ were compared between 97 LCP and 37 NFC and data from 42 pre-pandemic HC (Figure-2A). Missing data were due to shortage of serum/technical issues).

**Figure 2:**
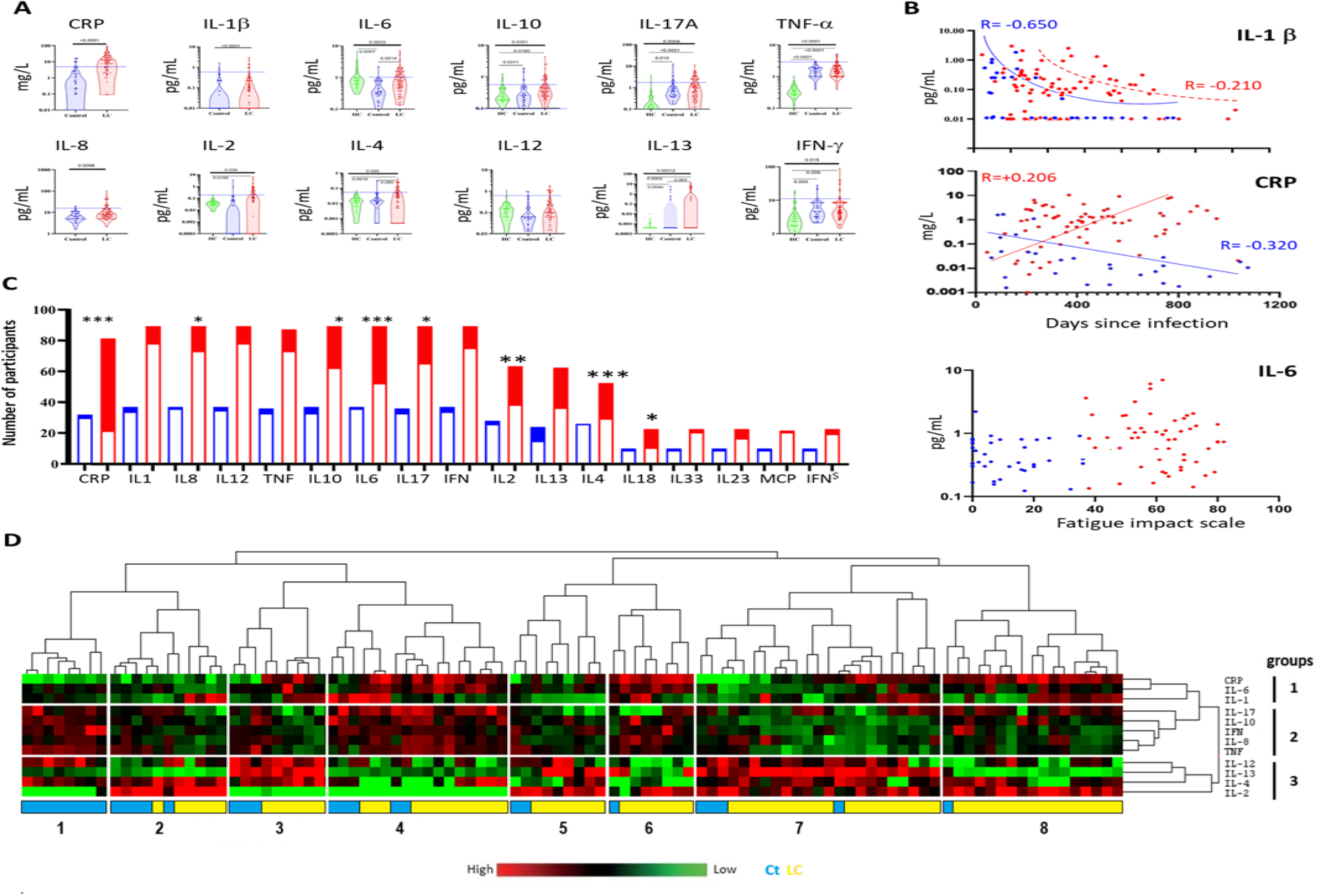
Systemic inflammation and circulating cytokines expression. Levels of CRP and cytokines were measured (using high sensitivity assays) in LC red, variable n between 32 and 97) and NFC (Bleu, n between 10 and 37) and pre-pandemic controls (n=42). **A)** Violin plots described levels. Distributions were highly skewed in LC whereas in NFC, fewer outliers were observed (not for all cytokines). ANOVA (thick lines) and MWU (thin lines) were used to test difference in levels. P-values are displayed on the plots. **B)** IL-1β and CRP levels reduced over time since infection in NFC but CRP increased in LC while IL-1β reduced with a marked delay (spearman correlation, rho-coefficient displayed). Other cytokines were not significantly impacted by time, while not for age, sex, or with respect to FIS as shown for IL-6. **C)** Cytokine levels were dichotomized based on whether they fell above the 95% confidence interval of the distribution in NFC, excluding outliers exceeding 2 standard deviations, with a cut-off marked by dotted blue lines. Of note, this cut-off was different from that of pre-pandemic data for TNF, IL-17A, but similar for other cytokines. Histograms of number of cases with raised values (filled) and within the range of values in NFC (open) are displayed with significance of the change in frequencies tested by chi^2^ [^$^ for IFN-α2, *** p<0.0001, ** p<0.001, * p<0.05]. **D)** Patterns of CRP/cytokines levels were analysed using CLsuter-3 (after log-transformation) and visualized on a heatmap of levels. CRP/cytokines segregated into 3 groups and patients into 8 clusters of patients (pink) and controls (blue).

CRP levels (distributed over 6 log range) were elevated in LCP (p=3.9x 10^-9^) and above the lower limit of normal (>5mg/L, Leeds area) in 60/82 (73%) LCP compared with 4/32 (12%) NFC (p=4.8x 10^-10^). Importantly CRP levels increase in LC over time since infection (rho=+0.206, p=0.040) but decreased in NFC (rho<-0.320, p=0.05).

Levels for IL-2, IL-4, and IL-10 were similar between pre-pandemic HC and NFC. In contrast, IL-13/IL-17A/TNF/IFN-γ showed higher levels in NFC (p<0.01). This was irrespective of the severity of the initial infection. Conversely, levels of IL-6 (p=0.0007) were lower in NFC compared to HC These discrepancies suggest the involvement of various inflammatory pathways in the resolution of the COVID-19 infection. Median cytokine levels were not significantly different between LCP and NFC except for IL-4 (p=0.05) and IL-6 (p=0.0014), despite highly skewed distributions. Analysis of cytokine levels with respect to demographics (age, sex, infection severity) showed no associations in LC or NFC (not shown).

With respect to time since infection, correlations suggested a reduction in IL-1β levels over time in NFC (Figure-2B, rho=-0.650, p=9×10⁻⁶), and a trend in LCP (rho=-0.210, p=0.020). Trend reductions were also observed only in NFC for IL-4 and IL-13 (rho<-0.325, p>0.030 and p=0.046) while IL-2 and TNF significantly increased (rho>+0.350, p>0.042 and p=0.0017). This was not observed in LC. Conversely, IFN-γ levels reduced with time in LCP (rho=-0.302, p=0.002) but were stable/low in NFC regardless of time. Therefore, most cytokines returned to “normal” shortly after infection (typically before 12 weeks), while this normalization does not occur in LC. There was no association between CRP/cytokines and FIS as illustrated for IL-6.

We dichotomised cytokine levels as normal or raised above data in NFC. This approach revealed significant changes in the frequencies of participants with raised cytokine levels (Figure-2C).

To investigate specific cytokine patterns, we employed the Cluster-3 algorithm (Figure-2D). This segregated CRP/cytokines into 3 main groups:

1. related to inflammation (CRP/IL-6/IL-1β)
2. with cytokines involved in Th1/Th17 polarisation and inflammatory/regulatory responses (IL-17A/IFN-γ/TNF/IL-10) or pain (IL-8)
3. Th1/Th2 cytokines (IL-12/IL-13/IL-4/IL-2)

This further delineated 8 participant clusters with CRP and IL-2 levels being particularly homogenous:

`o` one cluster (#1) consisting solely of NFC (n=8) defined by high levels of group-2 cytokines, regrouping most of the NFC-outliers for IL-17A/IFN-γ/TNF/IL-10/IL-8 while in the absence of CRP/IL-6/IL-1β.
`o` 3 clusters (#4, #6, #7) with high levels of either 1 or 2 of the cytokine groups
`o` 4 clusters (#2, #3, #5, #8) appeared more mixed but driven by low levels of one groups of cytokines. Demographic data across these clusters revealed no significant differences in age, sex, infection severity (2 asymptomatic patients/3 mild/2 moderate/1 severe) or FIS. Time since infection was not different between Cluster-1 and other NFC. Association with symptoms cluster are discussed further down.

### Chronic infection

One hypothesis proposed to explain LC is the persistence of live viral particles as opposed to a "sterile" disease with complete viral clearance. Here, we employed a surrogate measure for viral persistence by detecting the nucleocapsid (N) protein in serum. In LCP (Supp-Figure-S4), the N-protein was detected in 10/39 (26%) cases, with a median optical density (OD) of 2.45. It was also detected in 6/21 (28%) of NFC, although at lower levels (0.95 OD). The presence of the N-protein was neither correlated with the severity of the initial infection, FIS nor time since infection. Furthermore, no association was found with abdominal pain/nausea reported in the C19-YRS-PRO/PROM.

### Inflammasome

Given that we observed elevated levels of both IL-1β and IL-18 in LC, we investigated the potential activation of the NOD-like receptor family pyrin domain-containing 3 (NLRP3) inflammasome^[32]^. We measured levels of apoptosis-associated speck-like protein containing a CARD specks (ASC), a marker of NLRP3 inflammasome activation, in serum samples from 23 LCP and 12 NFC (Supp-Figure-S5). We detected increased levels of NLRP3-positive ASC specks in both NFC and LCP compared to pre-pandemic HC, but with no difference between them. Similar results were observed in the levels of NLRP3-negative ASC specks.

### Lymphocytes subsets

We investigated multiple lymphocyte subsets using flow cytometry. Due to constraints with amount of material, we performed a pilot lymphocyte phenotyping on 16 LCP and 11 NFC, using frozen PBMCs. Although large differences appeared in the frequencies of many sub-populations analysed (Supp-figure-S6), similar results in NFC and LCP were observed for lymphocytes lineages, B-cells and monocyte subsets. For NK and CD56+T-cells, the expression of 3 markers was different between groups, showing lower NK/CD16+ (p=0.042) and higher NK/CD62L+ (p=0.0007) and both lower CD56+T-cells/CD27+ (p=0.0075) and CD56+T-cell/CD62L+ (p=0.0011).

### T-cells exhaustion and differentiation

T-cell expressing high levels of exhaustion, senescence or activation markers were observed with high heterogeneity (PD-1/CD44/CD38/CD161/LAG3/TIM3/CTLA4), and affecting CD4+T-cells more profoundly than CD8+T-cells (Figure-3A, representative donor #1 and #2). Another panel addressed differences in naïve and memory T-cells (CD45RA/CD45RO) and their expression of several markers (CD27/CD62L/CCR7/CCR6/HLA-DR/CD57) with even more heterogeneity (donor #1, #2, #3).

**Figure 3:**
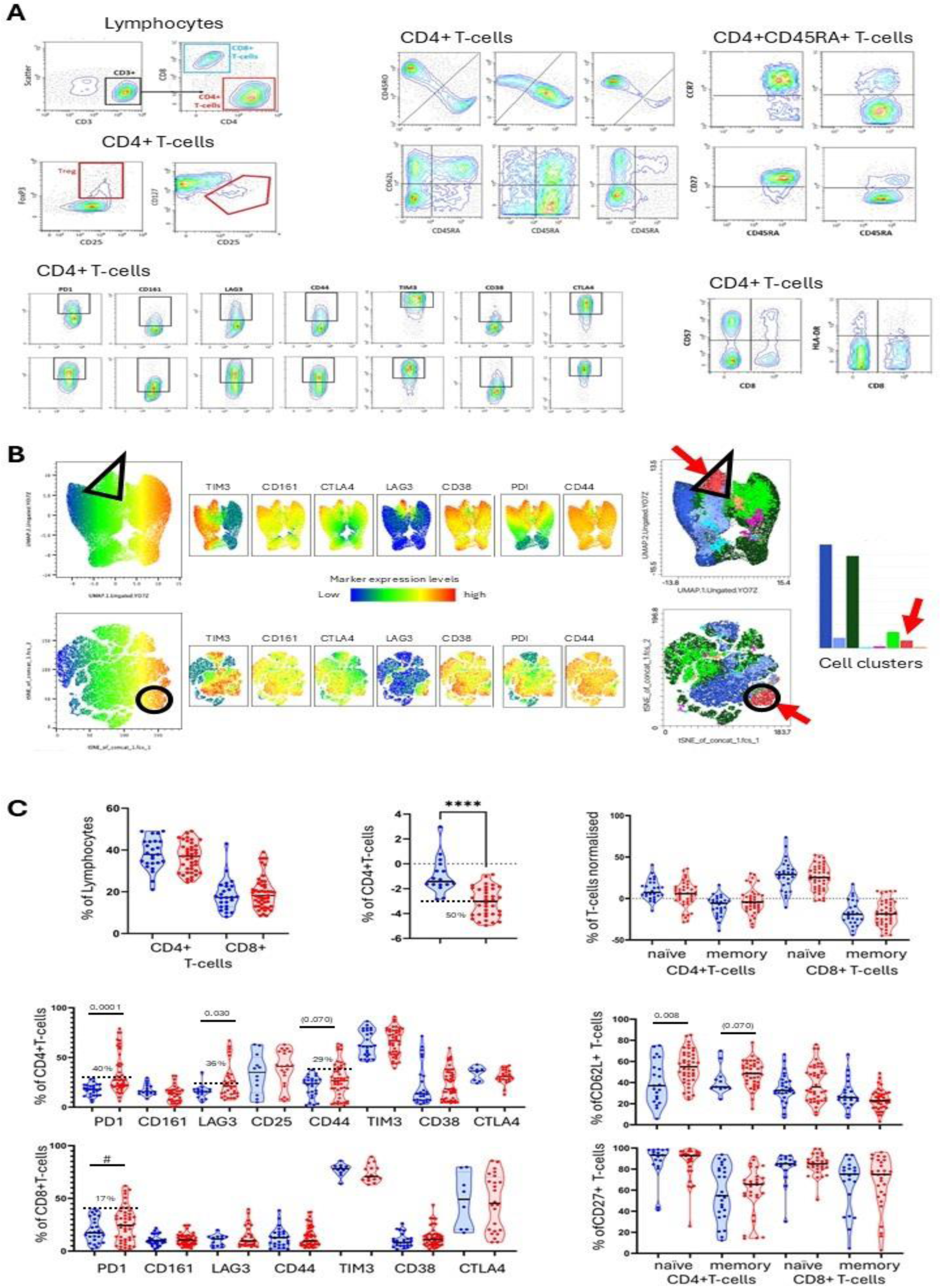
T-cell exhaustion, differentiation and Treg. **A)** Flow cytometry was used to analyse the expression of 17 markers on CD4+ and CD8+ T-cells (Lymphocyte panel) and Treg were identified as CD4+/CD25^high^/FoxP3^high^/CD127^low^. Most markers showed continuous expression rather than positive-negative population. Frequencies of cell in main/minor sub-population were variable and mean fluorescent index were not informative. Furthermore, profiles of expression were very variable for most markers as highlighted in representative donors on CD4+T-cells or naïve CD45RA+ CD4+T-cells (#1, #2, #3). Frequencies of cells expressing high levels of individual markers were recorded (as % of CD4+T-cells or % of naïve cells) using gates set using the contour plots for the main population (red-to-light green lines) versus minor population (green-to-blue lines). Multi-gating with up to 7 markers per panel was not attempted. **B)** Multi-marker analysis was performed for the panel including 7 exhaustion markers. Data from the 11 samples analysed the same day were concatenated and a tSNE and Umap analysis performed, followed by cell clustering using Flow-Jo. The cluster analysis suggested a subgroup of cells (arrow, triangle and circle) expressing high levels of the 7 markers however, not fully overlapping in all the cells in the triangled/circled area as pointed by the black arrows for some LAG3-, CD38-CD44-cells. **C)** Data were reported as % of CD4+ / CD8+ T-cells and plotted using violin plots (NFC blue, LC red). Naïve, memory and frequencies were normalised. Significant differences are highlighted (MWU, bars, p-value). Dichotomisation of levels raised above the distribution in NFC (cut-off dotted line) was performed and indicated by % of patients with raised frequencies for relevant markers. (**** p<0.0001, # p<0.100)

We performed a multi-marker analysis concatenating data for the panel including 7 exhaustion markers, using 11 LCP analysed the same day. This produced tSNE and Umap plots (Figure-3B) and identified 8 clusters of cells. One of the clusters (red-arrow, triangle and circle) included cells expressing high levels of the 7 markers, although not in all cells, notably for LAG3 (black-arrow).

Gating for positive-cells was performed for individual markers (Figure-3C as indicated on panel A, LCP n=47, NFC n=26). NFC showed relatively homogenous distribution of frequencies, while LCP showed more widely distributed data, suggesting disturbances of several markers (PD1+/LAG3+/CD44+/CD38+) significant on CD4+ cells for PD1+/LAG3+ and almost for CD44+. The same was seen in CD8+T-cells but less pronounced (trend for higher PD1+ and lower TIM3+). Following dichotomising data above the distribution of NFC values, the most frequently observed increase of positive-cells was for PD1+ (39% of cases), LAG3+ (32%) and CD44+ (25%) (only for PD1+ in CD8+T-cells (15%). In NFC, CD4+ memory cells were reduced (median - 4%), and even more in CD8+T-cells (−21%). Conversely, naïve cells were slightly above expected median in CD4+ (+5.5%), and in CD8+ (+32%). There was no difference between NFC and LCP, nonetheless showing a profoundly perturbed state of differentiation in CD8+T-cells, as could be expected following SARS-CoV2 infection. Naïve CD4+/CD62L+cells (but not CD8+) showed higher frequencies in LCP (p=0.008). Reduced frequencies of memory CD4+/CD57+ were also seen (Supp-Figure-S7, p=0.0066), and similar trends were consistently seen for naïve/memory subsets for both CD4+and CD8+. CCR7+ and HLA-DR+ frequencies were not altered, although a higher median was notice for naïve CD8+/HLA-DR+ in LCP.

We then investigated demographic parameters influencing cell subsets in both NFC and LCP. No difference was observed between sex. Spearman correlations were used however, limited by low number of data point (SUPP-Table S1, combined with data on B, NK, CD56+T-cells and monocytes). Nonetheless, relationships with age were observed in NFC but were lost in LCP (except one). Vice versa, associations in LC were not present in NFC, while some were in opposite directions. Similar results were obtained for correlations with FIS or time since infection. Association with clusters of symptoms are discussed below.

### Regulatory T-cells (Treg)

We quantified Treg populations (Figure-3A, CD25^high^/FoxP3^high^/CD127^low^) as previously reported^[33]^ and observed reduced Treg frequencies in LCP (Figure-3C, NFC n=16, LCP n=36, p=0.00008). In NFC, Treg reduced with time since infection (rho=-0.368, p=0.081) and increased with FIS (rho=+0.370, p=0.087). This pattern was no longer seen in LCP. Surprisingly, Treg frequencies were positively correlated with the PROM, fatigue/pain/mobility/breathing issue at rest (rho>+0.400, p<0.018).

### biomarkers of Phosphate mwtabolism

Many metabolic factors are implicated in fatigue and muscle weakness. Inorganic phosphate (Pi) is released from tissues during fatigue and interplays with inflammation^[34–36]^ In addition, the immune system regulates the production of the phosphaturic hormone fibroblast growth factor-23^[37, 38]^ notably via pro-inflammatory cytokines and lipocalin-2. We hypothesised that levels of phosphate and phosphate-regulating proteins could be altered in LCP. We measured circulating levels of Pi, LCN2 and FGF23. No significant difference in the distributions of data were observed, despite skewness in LCP (Figure 4A). Levels of Pi were increased in 23% of LCP (n=50, none of the 16 NFC), while high levels of LCN2 (16%) and intact-FGF23 (15%) were observed in LCP. In NFC, there was associations between low LCN2 or FGF23 with FIS while an increase in FGF-23 was observed with time since infection. None of these effects were observed in LCP suggesting disturbances in overall phosphataemia.

**Figure 4:**
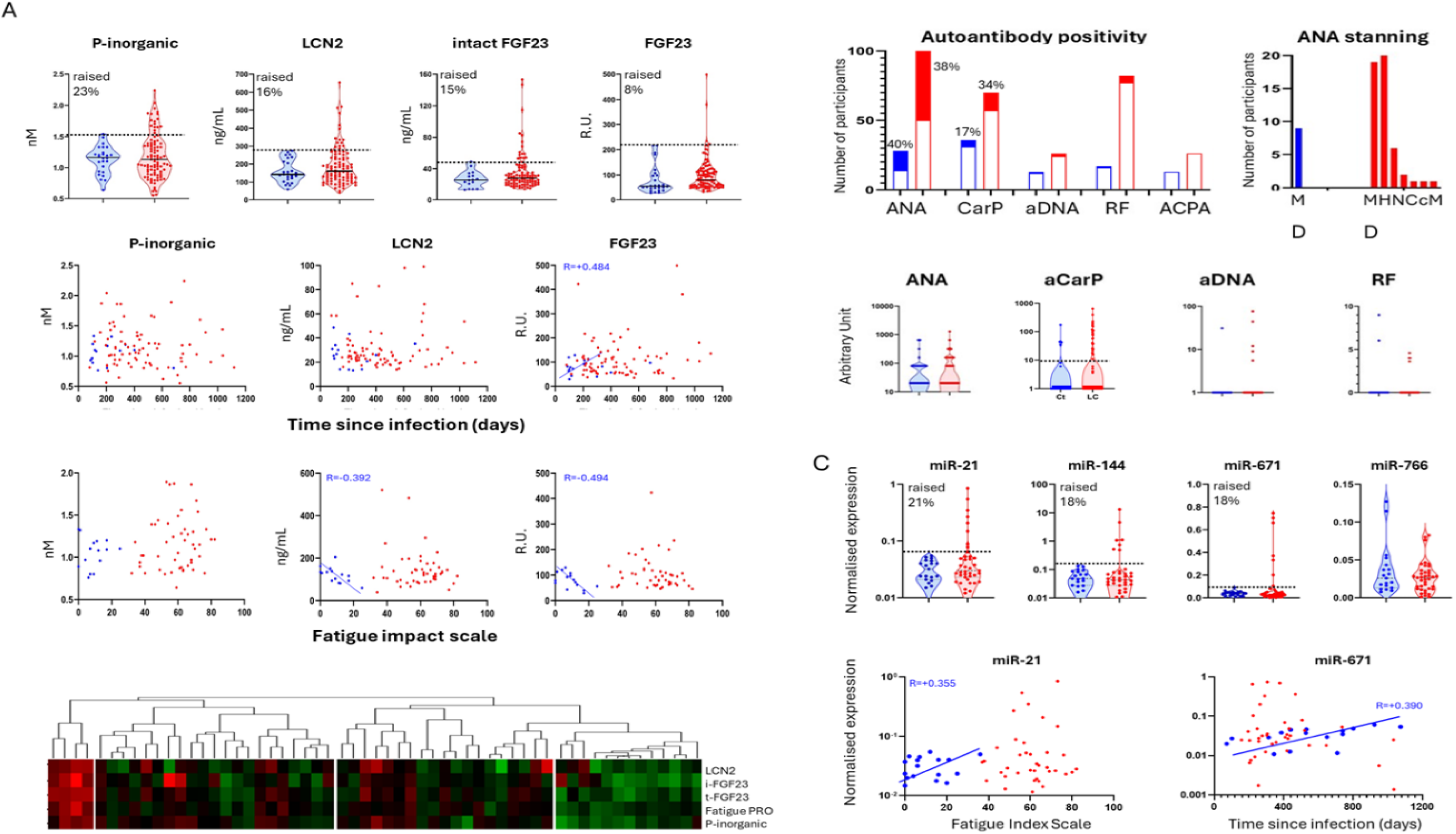
Phosphate metabolism, autoantibodies and microRNA. **A)** Phosphate inorganic and its regulator LCN2 and FGF23, were measure using commercial biochemistry assays in LC (red, n=50) and compared to NFC (blue, n=16-26 pending available serum/data). Levels were not statistically different but ∼20% of LC patients showed raised-levels (above the overall NFC distribution, no outlier). Relationships between time since infection or FIS with FGF23 and LCN2 were observed in NFC while absent in LC. Clustering of data with symptom-PRO/PROM in LC segregated 2 clusters of patients with very high and very low biomarkers/fatigue-PRO, comprising the most raised levels (outliers) and the lowest value for all biomarkers respectively, with another 2 clusters with mixed data. Other PRO/PROM did not associate with these biomarkers. **B)** Autoantibodies were detected with various ELISA tests and number and % of positive cases are presented as filled histograms (negative cases as open). No significant difference in frequencies were observe. ANA staining presented in a dotted (D) pattern for all NFC but was variable in LC with cases showing dotted, homogeneous (H), nuclear (N), cytoplasmic (C), centromeric (c) or mixed (M) pattens. Levels of antibodies were also heterogenous but not higher for ANA and anti-DNAor RF while higher at median levels for anti-CarP (p=0.101) in the samples with detectable levels (>10 AU/mL). **C)** The expression of 4 candidate miRNAs were measured in serum samples by qPCR in LC (n=39) and NFC (n=19). Again, levels of expression were very skewed in LC but not in NFC while not significantly different. Dichotomisation suggested raised-expression in ∼20% of LC cases. miR-21 was related to FIS and miR-671 to time since infection in NFC(rho displayed on graph), but not in LC.

### Autoantibodies

We confirmed the presence of ANA in 50% of both LCP (Figure-4B, LCP n=100, UK+France) and NFC (n=24). However, the intensity of ANA staining was notably higher in the LCP. Moreover, the ANA staining patterns differed between groups, with a dotted pattern consistently observed in NFC (100%), while LCP exhibited a broader range of patterns: dotted (38%), homogeneous (40%), nuclear (12%), and other atypical patterns (10%), including centrosomic, cytoplasmic, and membrane-associated staining. Anti-DNA antibody-positivity was seen in 1/13 (7.5%) NFC and in 5/77 (6.5%) LC. Persistent inflammation can also trigger post-translational modification (PTM) of proteins creating neo-epitopes. Carbamylation of protein is associated with stress and anti-carbamylated protein autoantibodies (anti-CarP) were observed in ∼50% of hospitalized cases of acute COVID-19, also associated with Rheumatoid Factor (RF) in 25% of cases^[39, 40]^. RF-positivity was identified in only 5/40 participants, suggesting this response may be transient—particularly considering that most participants were not hospitalized. Anti-CarP antibody reactivity was clearly detected above the test background in a 35/107 samples while raised in 4 (<10 AU/mL) but below detection in 70 samples. Of these35 samples, 5/19 (26%, median 48 AU/mL) were NFC and 31/88 (34%, median 106 AU/mL) were LCP, which was not statistically different (p=0.101). No correlation was observed between levels of anti-CarP reactivity and severity of the infection, time since the infection or demographic variables. Antibodies against citrullinated proteins (a PTM commonly associated with inflammatory joint diseases), were not detected in any of the participants tested.

### miRNAs candidates

We performed a bibliographic search of the literature for miRNAs candidates associated with fatigue, T-cell exhaustion, inflammation, autoimmunity, mitochondrial activity or neurological dysfunctions to select candidates (Supp-figure-S8). We selected 4 candidate miRNAs for testing (miR-21, miR-144, miR-671, miR-766). RT-qPCR was performed on total-RNA extracted from serum in LCP (n=39) and NFC (n=19). Three miRNAs showed patterns similar to other biomarkers (Figure-4C), with raised levels in 10-15% of LCP, while not significant at expression levels. The expression miR-21 and miR671 correlated well with each other (rho=0.790, p=5.6×10^-7^) while not with miR-766. In Addition, miR-21 showed trend correlation with FIS in NFC (rho=0.355, p=0.070) and miR-671 with time since infection (rho=+0.390, p=0.065), which were both lost in LCP.

### Multi-marker integration

We investigated immunological and non-immunological candidate biomarkers associated with some of the hypothetical mechanisms proposed for LC (5 demographic, 28 serum tests, 80 cell subsets, 16 PRO/PROM, 1 overall health score). A matrix of correlations between serum and cell subset biomarker levels in LCP (only when n>8) is provided in SUPP-Figure-S9/Supp-Excel file showing many weak associations suggesting collinearity between all features measured while no evidence of highly associated biomarker (as n=8 for most of the rho>0.600). In the pilot phase, multiple platforms were used. Samples in the 2^nd^ stage, were tested for the candidates/platform suggesting differences due to limitations in material/funding. Altogether, comparing NFC/LCP, levels of 20/108 biomarkers tested were significantly different (Supp-Figure-S10, 15 with p<0.001), and 5 more showing trends). Dichotomised data for 25 biomarkers allowed to select 3 more candidates. Levels of these 28 biomarkers (and 41 biomarkers performed on the same platform) confirmed correlation with similar and more importantly opposing relationships with FIS and duration since infection in NFC versus LCP.

### Overlap between possible disease mechanisms

Using dichotomised data for the biomarker that showed significant departure from data in NFC, raised-CRP was the most frequent feature (73% cases), while most of the other biomarkers tested were associated with 15-50% of cases. We assembled biomarkers in various groups (raised-CRP, cytokines, anti-CarP+, phosphate metabolism; although not enough cases could be used with miRNAs data, loss of Treg, CD4+ subsets and CD8+/PD1+). We first used Venn diagrams to visualise overlap between features (Figure-5A). For Serum candidates, 10% of patients (6/62) showed disturbance in all biomarkers suggesting an important overlap between potential mechanisms. Inflammation was however, also seen alone (11/62) while rarely for other features (raised cytokine, anti-CarP or phosphataemia markers, 3/62). Most patients displayed raised-CRP and at least 1 more feature (39/62). A similar approach for cell subsets showed that both Treg-loss and CD4-exhaustion were observed alone (5/32 and 6/32) as well as with inflammation (5/32) but inflammation was not observed in the absence of T-cell abnormalities. LCP without feature were the same in both analyses.

**Figure 5:**
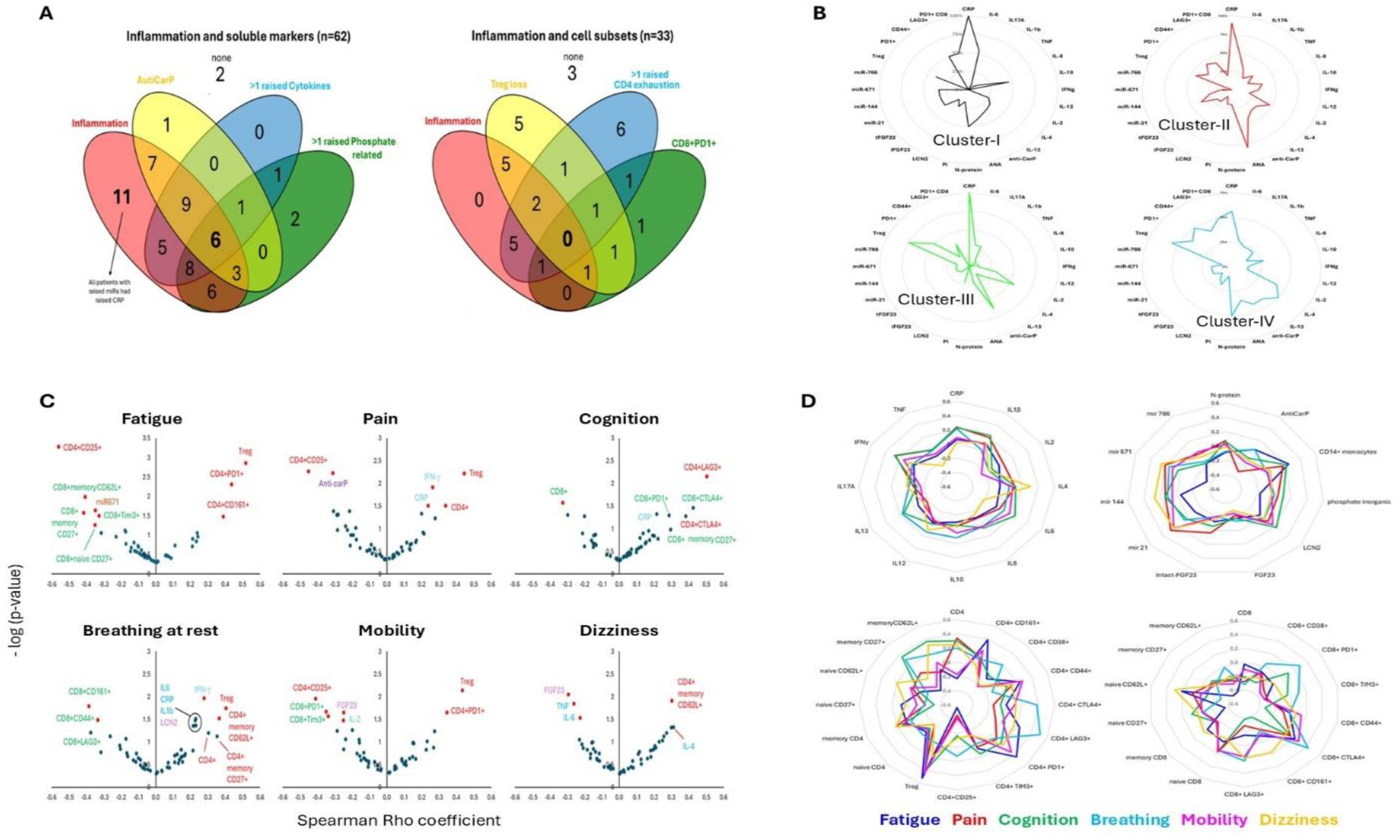
Biomarker overlap and association with symptoms. A) We used dichotomised data for 28 variables (selected from the soluble markers and cells subsets tested with significant association with LC as described in the manuscript), combining raised-CRP (yes/no) with, on one hand (i) cytokines (>1 cytokine with raised level), (ii) presence of anti-CarP (iii) alteration in Pi metabolism (>1 marker with raised level) for soluble biomarkers (62 cases) and on the other, with (i) loss of Treg, (ii) >1 exhaustion marker raised on CD4+T-cells and (iii) raised CD8+PD1+ for cells subsets (33 cases). Overlaps between biomarker categories are represented using Venn diagrams. B) We looked at the frequencies of raised data for the 28 biomarkers separating the patients based on Cluster defined by symptom PRO/PROM in Figure-1. Number of cases available for each biomarker was variable but the lowest number considered was 7. For examples, CRP was raised in 100% of patients in Cluster-I and III while only in 60% of patients in Cluster-IV. Treg were more frequently lost in Cluster-III and IV. ANA were most frequent in Cluster-II (80%) and anti-CarP in Cluster-II (65%). C) Volcano plots representing the strength of the correlation (Spearman rho coefficient) versus −Log_10_(p-value) are described for 53 potential biomarkers with data for >25 patients (no other selection criteria) and 3 symptoms PRO/PROM selected by patients (fatigue pain cognition) in the C19-YRS questionnaire as well as with breathing (at rest), mobility and dizziness. Biomarker dots in red have significant unadjusted p-value (p<0.05) and rho> 0.300 while blue dots may have good rho values but non-significant p-values. CD4+T-cell biomarker are highlighted in red, CD8+ in green, CRP/cytokines in blue, miRNAs in brown and markers of phosphate metabolism in pink. D) Radar plots representing the strength of the associations (rho-coefficient) between the same 53 biomarkers and the 6 chosen symptoms PRO/PROM. This highlights particular candidates associated with each symptom, for example dizziness (yellow line) and IL-4. Altogether it also suggests that CRP/cytokines are all similarly associated with the 6 PRO/PROM as well as other soluble biomarker, with the exception of the miRNAs. In contrast, there was much more heterogeneity between symptoms’ association with specific CD4+T-cell subsets (highly positive/negative rho), also seen for CD8+T-cells.

We then looked at these biomarkers (dichotomised) with respect to the 4 clusters of patients identified in Figure-1. These presented very different patterns with high/low frequencies (Figure-5B, % of cases with raised-data).

- <u>Cluster-I</u> including severe breathing issues and high number of other symptoms had high frequency of raised-CRP (100%), IL-6/IL-10, N-protein (>50%), miR-21/144/671 (33%) and FGF23 (33%), excluding IL1β/TNF/IL-17A and exhaustion markers (<8%), suggesting inflammatory pathways at work in that cluster:
- <u>Cluster-II</u> including breathing issues + depression, had raised-CRP (90%), ANA (80%), other biomarker being present in with no noticeable enrichment/exclusion.
- <u>Cluster-III</u>, including severe fatigue/pain/cognition/sleep and mobility showed raised-CRP (100%), Treg-loss (75%), anti-CarP (62%), IL-2 (55%), CD4+PD1+ (50%), excluding TNF/IL-1β/IL-10 (not IL-17A) and phosphatemia (all <10%), and CD8+PD1+and N-protein (0%).
- <u>Cluster-IV</u> for fatigue/pain/cognition/sleep and Anxiety/depression/Dizziness had raised-CRP, IL-4, ANA, Treg-loss, CD4+CD44+, CD8+PD1+ (all about 50%), excluding phosphatemia and miRNAs (<10%).

### Correlation with symptoms (PRO/PROM)

We integrated biomarkers data with PRO/PROM. Correlations were used to investigate potential biomarker associations with 6 PROs (Figure-5C/D volcano and radar plots), limiting analysis to the 69 variables with data in >25 LCP. Biomarkers were differentially associated with specific symptoms. Some had unique profile, such as CRP only related to cognitive issues (despite being high in 75% of patients, IL-4 with dizziness, autoantibodies or miRNAs with pain). In contrast, Treg, CD4+CD25+ and CD4+PD1+ cells associated with multiple symptoms, CRP only with pain/cognition, several cytokines with pain/breathing. Several subsets of CD4+ and CD8+ cells associated with multiple symptoms although often negatively for CD8+ and positively for CD4+.

### Possible biomarkers discriminating LCP from NFC

We attempted to use the integrated multi-platform data-system for mathematical modelling towards developing tools for diagnosing, monitoring and stratifying patients. Missingness was a major issue, even starting from 69 variables with data in >15 NFC and >25 LCP. Multi-variate analysis could not be performed on the overall dataset, imputation being overfitting and missingness not at random. We therefore reduced the number on biomarkers to the most likely candidates to perform missing data imputation on a limited number of variables. Using univariate analysis, the 28 candidates identified above were selected and data imputed into 2 datasets, separating serum candidates (LC n=87, NFC n=40) and cell subsets (LC n=47, NFC n=22). Regression analysis suggested CRP (yes/no) and IL-8 (levels) to discriminate LCP from NFC (Supp-Table-S2 accuracy 79.5%) and Treg, CD4+PD1+, CD4+CD161+ (accuracy 88.5%). Combining participants with both datasets (LCP n=41, NFC n=19), the model selected only the 3 subsets.

### Biomarkers associated with PRO-change over time

C19-YRS-PRO/PROM data were available at 6-12 months (n=65). Based on discussion with our LCP, we selected the 3 symptoms they wish the most to reduce: fatigue/pain/cognition. We looked at reduction/increased in PRO scores and associated an upward or downward change in patients with clear change (Figure-6A, >3 points change in 1 PRO or >1 point change in all 3 PROs) towards improvement (12/65) or deterioration (9/65). Most patients however, showed limited changes, mainly in 1 Pro only (Figure-6A). None of the 4 cluster showed any association with either outcome (SUPP-Figure-S11).

**Figure 6:**
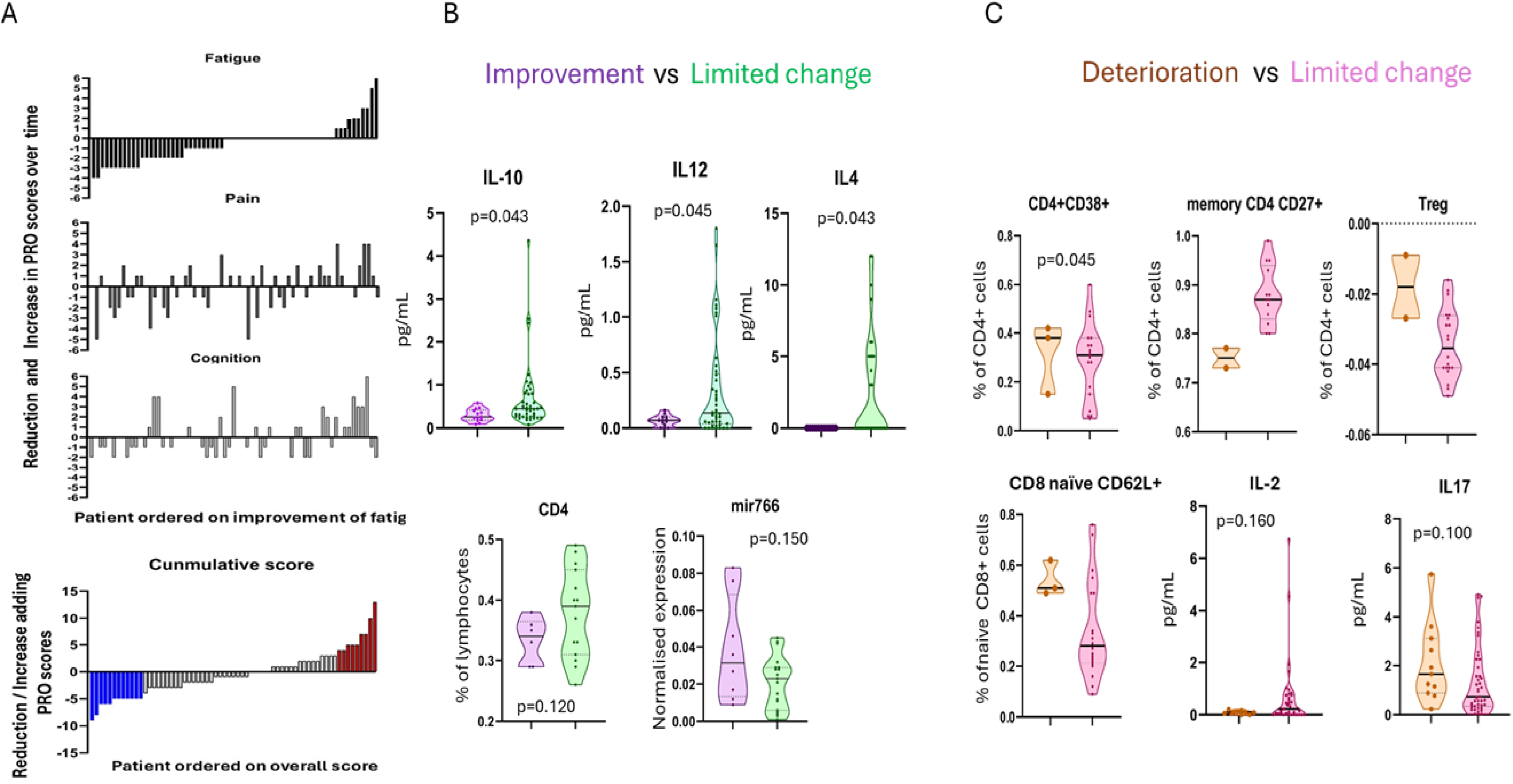
Biomarker and change in PRO/PROM over time. **A)** Change in PRO/PROM scores (data at 12m – data at time of blood collection) were calculated for fatigue pain and cognition in 65 patients. Changes range from −4 (reduction of severity) to +6 (aggravation). Ordering patients on change in fatigue scores, showed that change in one PRO/PROM is not always associated with change in the others. Adding the 3 changes to create a cumulative score (bottom panel) and re-ordering patients allowed to define a group of 13 patients with a cumulative score of −5 and less (blue) suggesting overall reduction in symptom severity and a group of 13 patients (red) with increased severity (+5 and more). Most patient however, showed limited PRO/PROM change. **B)** To investigate biomarkers with respect to improvement or deterioration of symptom, we selected patients with clearest changes with either >3 points change in 1 PRO/PROM or >1 point change in all 3 PROs all in the same direction towards improvement or deterioration. This allowed us to select a group of 12/65 patients with improvement and 9/65 with deterioration. IL-10/IL-4/IL-12 were lower in LCP improving (p<0.05) compared to LCP with no change (green) and CD4+T-cells and miR-766 could be relevant, although not reaching statistical significance. In patients with aggravation (orange, n<5 cases for most biomarkers), Treg, and CD4/CD8 subset of cells showed trends, with IL-2 (p=0.032) and possibly IL-17A.

We attempted analysis of biomarkers with improvement or deterioration. Case numbers were too low (n<5) for many biomarkers allowing only indicative comparisons. IL-10/IL-4/IL-12 were lower in LCP improving (Figure-6B, purple p<0.05) compared to LCP with no change (green) and CD4+T-cells and miR-766 could be relevant. In patients with aggravation (Figure-6C, orange, n=3 cases for most), Treg, and CD4/CD8 subsets showed trends, with IL-2 (p=0.032) and IL-17A.

## DISCUSSION

In this study, we evaluated candidate biomarkers rationalised based on disease mechanisms hypothesised to contribute to LC. Multiple platforms were used suggesting strong candidates but also eliminating some, associated with various mechanisms involved in LC.

**Persistent inflammation** is the most frequently observed feature of LCP (73% cases), increasing over time while reducing in NFC. Cognitive issues were associated with CRP and pain with CRP/IL-6. CRP is a well-established biomarker of systemic inflammation, though to contribute to cognitive decline by disrupting the blood-brain barrier (BBB)^[41]^. Cognitive decline and dementia have been linked with CRP in a complex relationship depending on age and sex^[42–46]^ and not necessarily a direct cause, other factors (genetics, lifestyle, pre-existing conditions) playing a role. IL-6 may be more directly mechanistically relevant^[47, 48]^ while as a biomarker, CRP may be more sensitive/specific hence associated with the PRO/PROM^[49]^. Several mechanisms were explored to explain where inflammation comes from and why it does not subside. Persistence of the virus or lingering antigens was proposed^[50, 51]^. This hypothesis stemmed from the dissemination of live/replicating viral particles in various body niches during acute infection^[52, 53]^, leading to incomplete viral clearance^[54]^. Viral shedding in stool samples lasted at least 4 weeks in 95% of severe/hospitalised COVID-19 patients (who subsequently fully recovered), while after 7 months only 4% were positive^[55, 56]^. **N-protein** was present in both NFC and LCP, associated with high TNF/miR-144 and low miR-766, but not with CRP/exhaustion or with IFN-γ which could have been expected from previous reports^[57]^. Abdominal pain/nausea and vomiting associated with SARS-COV-2 in the gut^[55, 56]^, were not reported by N-protein+ patients. While N-protein does not indicate whether fully replicating viral particles are present, our data argue for a non-specific occurrence. Our data also suggest the NLRP3-pathway is not involved in persistent inflammation, which also aligns with the normalisation of IL-1β levels over time in both groups. Another proposed origin is virus-induced dysbiosis resulting in increased gut permeability^[58-61]^ which we did not address. Dysbiosis would also result in metabolic perturbations^[62, 63]^ with reduced production of immunoregulatory metabolites (particularly small chain fatty acids) regulating the generation of inducible-Treg or the inhibition of Th17^[64–68]^. In view of our data on Treg loss and high IL-17A, it would be of great interest to measure both gut permeability and gut metabolic output in LCP.

### Cytokines

A cytokine storm has been described^[69]^ resulting from overactivation of immune system^[70]^, notably observed in severe COVID-19. More detailed exploration of symptoms during infection may help clarify whether LCP with breathing issues, may be associated with a cytokine-mediated mechanism. Tissue damage resulting from this storm with multi-organ failures and release of various molecules (PAMPs, DAMPs or cell death activators), initially provided rational for LCP to develop from severe COVID-19 cases, since refuted as most LC cases develop from mild/moderate (even asymptomatic) COVID-19. IFN-γ/IL-1β/IL-6 were related to pain and IL-4 to dizziness. Cytokines can directly affect the nervous system and are well known to contribute to pain, particularly in inflammatory conditions^[71–74]^. IL-4 can promote fibrosis in the heart^[75, 76]^, contributing to heart failure/hypertension, lung vascular inflammation and remodelling^[77]^, while IL-4 also have a protective role reducing blood pressure in experimental models^[78]^.

### Exhaustion

An increased proportion of exhausted CD4+T-cells was clearly observed for individual markers (up to 40% of cells) with sub-populations expressing multiple markers. Exhaustion is observed in the absence of inflammation as frequently as in its presence, suggesting that inflammation may not be its only cause. In Cluster-I, PD1+/CD44+ cells are not raised leaving way for the LAG3+ pathway, which is associated with chronic antigen exposure^[79]^, and Cluster-I is indeed enriched in N-protein+ cases. miRNAs candidates were also frequently disturbed in Cluster-I and they regulate PD-1 and TIM3^[80–84]^. Exhaustion promotes cytokine inhibition and low frequencies of raised IL-1β/IL-12/IL-17A/IFN-γ/TNF were seen in Cluster-I. In Cluster-II, raised-PD1 (33%) was seen with raised-miR-21 (37%) and their relationship is well known (Supp-data on miRNAs selection), while in cluster-III and IV, exhaustion is frequent (PD1+/CD44+/LAG3+) but with no miRNA disturbance. Therefore, various means to induce exhaustion appears possible while if is relatively common to all clusters. Perturbation of co-stimulatory signals is another cause of exhaustion, via epigenetic remodelling^[85, 86]^. We did not address co-stimulation on antigen presenting cells, while CTLA4+ cells showed lower median CD4+/CTLA4+. These appear to be important avenues to explore in future work towards understanding how these mechanisms may be responsible for the CD4+T-cell exhaustion in LCP but it clearly suggests that causes and consequences of T-cells exhaustion may not be the same in these 4 clusters. T-cell exhaustion is increasingly linked to cognitive issues (Alzheimer and neurodegenerative conditions^[87]^) with the presence of exhausted T-cell infiltrating the brain^[88, 89]^, due to disruption of the BBB. The cognition PRO was associated with exhaustion subsets (both CD4+ and CD8+) and exhaustion was also frequently seen in Cluster-III and IV with most severe cognition issues. Understanding the mechanisms underlying these observations remains to be elucidated while developing strategies to restore T-cell function could offer promising avenues for treating the cognitive aspects of LC.

### Treg

Treg was a highly specific biomarker for LC surprisingly associated with 4 PRO/PROM. Fatigue has a complex relationship with Tregs, some studies suggesting difference in frequency and others dysfunction^[90-93]^, depending on the diseases (chronic inflammation, autoimmunity, Treg/Th17 balance). Our data showing the reduction over time since infection in NFC suggest resolution of a Treg response as a hallmark of returning to health. Treg-loss was observed in correlation with raised-IL-17A suggesting that this imbalance may have an important role in LC. Cluster-III, including the most severe cases of LC is associated with Treg-loss (75%) and raised IL-17A (30%). PD1+Treg were observed in COVID-19^[94]^. This suggest an alternative where Treg cells may be exhausted in LCP, which may then explain their relationship with fatigue-PRO as observed in Custer-II and IV, while excluded in Cluster-I (no raised-PD1/IL-17A).

### Phosphataemia

Many metabolic factors are implicated in fatigue and muscle weakness (ATP/ADP, hydrogen ions, lactate, Pi, oxygenation, cortisol, IL-6) as regulators of the force and power of muscle contractile responses^[95]^. The availability of iron (Fe), Pi and phospholipids are the primary upstream regulators of the ATP/ADP balance. A such Pi is considered a biomarker of fatigue^[34]^,while factors such as LCN2 and FGF23 are responsible for the regulation of Pi and/or Fe^[37, 38]^. LCN2 is released from the liver in response to stress/anxiety^[96]^, it is involved in neuroinflammation anxiety, and cognitive function and age-related diseases of the central nervous system^[97–99]^. FGF23 regulations are essential for musculoskeletal functions (mobility) yet the spectrum of consequences for FGF23 excess spans beyond, with systemic effects, including cardiovascular, renal functions. Dysregulation of phosphataemia markers was individually associated with breathing/mobility/depression/dizziness. Abnormal levels of FGF23, leading to imbalance of the phosphatemia were associated with dizziness as a side effect^[100]^ and with impaired walking/mobility due to muscle weakness and bone pain^[101, 102]^. FGF-23 is also elevated in hypertension and can directly affect the heart by targeting myocardium cells^[103–105]^. Mobility is particularly affected in 2 Clusters, with both raised-LCN2/FGF23 in cluster-I and Pi more frequently raised in cluster-III while excluding LCN2/FGF23. Altogether, there appears to be an important role for the dysregulation of phosphatemia in LC.

### Autoimmunity

Thus far, studies on the presence of autoantibodies in LCP have focused on candidate antigens (anti-ACE2, MDA5CD255, SS-B/LA and more), some observed in severe primary infection^[39, 106]^. However, ANA were also present in post-infection controls, arguing for a potential artefact^[107]^. Our data are aligned with these reports^[39, 106]^. We observed an enrichment in ANA-positivity in symptom Cluster-II where breathing issue and depression are most frequent. ANA are linked with lung complications (pleurisy, fibrosis, cough, pain) and a relationship between ANA and depression also exists^[108]^. In contrast, we observed more LC-specificity for anti-CarP reactivity which were also observed in hospitalised COVID-19 patients^[39]^. Levels of anti-CarP antibodies observed during the acute phase of COVID-19 are higher than observed in LCP. Anti-CarP were also exclusively detected in the presence of inflammation. Carbamylation of protein (a non-enzymatic post-translational modification) originate from a stress reaction (isocyanic acid/elevated urea levels^[109, 110]^). The development of these autoantibodies may therefore be related to inflammation in LC but this does not explain why some controls develop anti-CarP. Levels of anti-Carp reactivity were associated with pain and most frequent in Cluster-III (highest pain). Autoantibodies, arising from various injuries (nerve damage, changes in nerve function) are often associated with pain via a Fc-region dependent excitation of nociceptors, while recently another hypothesis suggested that anti-nociceptors autoantibody binding can also directly cause pain^[111, 112]^. Carbamylation itself and anti-CarP antibodies are associated with join pain^[109, 110]^ as well as impaired healing^[113]^ and atherosclerosis^[114]^. Our data therefore suggest a degree of association between tissue damage and ANA/anti-CarP while unbiased approaches detecting novel reactivity (auto-antigen arrays) needs to be taken to determine whether true LC-specific autoimmunity contribute to the disease.

### miRNAs

miR-21 is known to correlated with CRP and IL-6 not surprisingly considering its role on modulating inflammation^[115]^, both for its resolution^[116, 117]^ or promotion^[118]^. As such miR-21 and miR-671/144 were also most frequently raised in Cluster-I, with raised-CRP/IL-6. miR-671 appears to be associated with atherosclerosis, rheumatoid or osteo arthritis, Crohn’s disease^[119, 120]^ with a prominent role in bone-cartilage hence possibly explaining mobility issues in Cluster-I. miR-144 is known as a stress-response microRNA^[121, 122]^, implicated in various biological processes^[123]^, with a biomarker role in depressive disorder and ME (severity of post-exertional malaise)^[124–126]^ and we observed a correlation with dizziness. miR-144 levels of expression were not correlated with the other miRNAs which may suggest a different origine. Our observations therefore suggest that miRNAs represent strong candidate biomarkers that may be used to identify further pathways to explore in LC (oxidative stress, ketamine, and more)^[121]^.

It is important to note that a definitive set of biomarkers for LC remains to be established. Using multiple hypothesis-led approaches suggested strong candidates by selecting features related to putative mechanisms, and yielded results even if we are cautious in their interpretation due to small numbers and the use of association (spearman or chi^2^). Several biomarkers showed association with LC (compared to controls) or symptoms/outcome which remain to be confirmed, and showed clear physiological relevance to possible mechanisms. The regressions performed are still only indicative and suggest that cell subsets are more predictive of LC than serum biomarkers. However, this is further limited by the characteristics of the groups, being post-diagnosis (even using NFC) which may be better suited for speculation about mechanism(s) of disease than for diagnostic biomarker research. Despite remaining highly speculative, the biomarkers associated with possible outcomes were very different between classification, improvement and aggravation of symptoms. A meta-analysis of >100 potential biomarkers (including cytokines/chemokines, biochemicals, vascular markers, neurological markers, acute phase proteins but not immune cell subsets)^[127]^, suggested 3 candidates (CRP, IL-6, TNF), by comparing both pre-pandemic HC and NFC, associating CRP with most symptoms. Most of these 28 studies had samples size varying between 10-300 but several analysed samples as early as 10 days after COVID-19, while most were taken before/over 12 weeks. Raised-CRP is definitely an indication of LC, and we further identified additional raised-cytokines in LC (IL-2/IL-4/IL-6/IL-8/IL-17A) while only weakly for IFN-γ/TNF/IL-13/IL-1β. IL-8 was observed in one of these studies with a similar recruitment to ours^[128]^ and IL-17A in another^[129]^. This does not however, allow further alignment between dataset, particularly considering that decline/normalisation of several biomarker levels in NFC had not yet occurred, which would prevent specific findings. Using a preselected proteome platform (OLINK, >2000 analytes) and 21-colour flow-cytometry ^[130]^, Gao *& Col*. observed skewed upregulation of many proteins in LC plasma compared to NFC reporting higher IL-18 (not significantly using platform statistics) but not IL-6, TNF or even CRP. Ten common markers in our study (tested in serum which may contribute to the discrepancies) showed raised levels in LCP. Despite using similar markers, our flow-cytometry approaches were too different to allow direct comparison, but some conclusion were similar with respect to monocytes, B-cells and cytolytic-cells and an increase in CD8+PD1+cells^[130]^, while we observed additional disturbances in CD4+ T-cells and NK-cell markers.

Our most important findings however relate to the multiple patterns observed and the overlap (or exclusion) of some features, defining clusters of patients within a syndrome rather than a unique disease (still taken with caution due to small numbers). Autoimmunity, raised cytokines and phosphataemia develop mostly in the context of inflammation (55/62 cases). In contrast, raised-CRP is only observed with CD4+T-cell exhaustion in half the cases (14/32) and miRNAs were dysregulated irrespective of CRP levels. On the other hand, persistence of viral antigen (free or encapsulated in extra cellular vesicle as recently described^[131]^), a role for the IL-1β-NLRP3 pathway or for ANA seem unlikely, while we have not ruled-out live virus hiding in tissue reservoir and/or persistence of gut dysbiosis. Our findings linking possible molecular pathway with LC clusters of symptoms related to these biomarkers, provided targets that could be addressed therapeutically with various types of interventions once further validated. Indeed, existing and novel therapeutic cytokine blockade should be considered, adjusted to patient profile as a precision medicine strategy. Patients with profiles falling in cluster-I would benefit of anti-IL6, rather than anti-IL1β and anti-IL17A for example. Restoring functionalities in exhausted cells could also help (targeting PD1/TIM3/LAG3^[132]^, rather than CTLA4). In view of the possible protective effect of elevated IL-2 (and conflicting data in COVID19^[133]^, combination therapy with bispecific-PD-1-IL-2s (effective on both CD4+/CD8+T-cells^[134–136]^) may help restore PD1+ exhausted (Cluster-I/IV) and Treg function (Cluster-III). Combination of dietary intervention and neutralizing antibodies (targeting FGF23 and LCN2)^[137]^ and LCN2)^[138]^ may also work in patients with elevated concentrations of FGF23 and disturbed phosphatemia (cluster-I). Finally, antagomirs blocking miR21^[139]^ (raised in Cluster-I/II) have been proposed as a therapeutic anti-inflammation strategy in many situations^[140–143]^. Since miR-21 also contributes to breaking the BBB^[144]^ and is implicated in pain^[145–147]^ (showing promises in pain models^[148]^), which is dominant in Cluster-III/IV, antoagomiR-21 could possibly be considered as a more universal option.

## Supporting information

submitted manusrcipt

## Data Availability

All data produced in the present study are available upon reasonable request to the authors

## Acknowledgements

The authors would like to acknowledge the kind help of Eva Maria Stork, LUMC, Leiden, The Netherlands in measuring ACPA. Kevin Guillemin and Anna-Emmanuelle Berger for their valuable technical assistance in blood sample collection/handling and ANA testing in Saint-Etienne.

## Supplementary material

### SUPPLEMENTARY METHODS

#### Patient recruitment strategy in UK

The National Health Service (NHS) LC pathway in Leeds, established in September 2020, is a community-based network of rehabilitation health centres across the city. Following referral from general practice, patients undergo a comprehensive triage assessment to confirm LC diagnosis and complete a questionnaire that we specifically designed for LC (the YC-19) ^[149]^, which includes pre-pandemic medical history. This captures core LC symptoms and provides a tailored online tool for evaluating severity, disability and general health ^[150]^. Subgroups of symptoms were created to analyse information across specific domains: Symptom Severity (SS), Functional Disability (FD), and Overall Health (OH) ^[151, 152]^.

Patients interested in participating in research were invited via email during their triage assessment to attend a visit at the University of Leeds (or at home) and to provide a blood sample (Figure 1a). Inclusion criteria were referral to the LC pathway, age 18-65, persistent fatigue, cognitive impairment and/or any form of pain, with allowance for other LC symptoms. Proof of prior SARS-CoV-2 infection was required, although patients infected before the availability of PCR testing (March-June 2020) were allowed. Demographic data and the Fatigue Impact Scale (FIS) (https://www.sralab.org/sites/default/files/2017-06/mfis.pdf) were recorded on the visit day. Access to electronic records of YC-19 questionnaire data was also granted.

Individuals who had fully recovered from COVID-19 without experiencing long-term fatigue were recruited as a no-fatigue control group (NoF-Ct) from university staff, invited through general email circulation. They provided demographic data and completed the FIS.

#### Blood samples processing

From participants in Leeds, 50 mL of heparinized blood and 9 mL of clotted blood were collected. Serum was separated from the clotted blood by centrifugation, performed within one hour of collection, and then frozen at −80°C. Peripheral blood mononuclear cells (PBMCs) were isolated from the heparinized blood by Lymphoprep density gradient centrifugation and cryopreserved in liquid nitrogen with 10% dimethyl sulfoxide (DMSO) in foetal calf serum (FCS). Heparinized plasma was recovered from the separation procedure and frozen at −80°C. In CHU-Saint Etienne, 9 mL clotted blood was collected and serum processed as above. Furthermore, data from 44 Healthy controls (HC) serum pre-pandemic were made available from Dublin.

#### Autoantibody measurements

##### RF

Self-coated 384-well plates were used, where human IgG was diluted to 10 µg/mL in Coating Buffer (pH 9.6) and coated onto the plates overnight at room temperature. Between each sequential step, plates were washed three times using Phosphate Buffered Saline (PBS) with 0.05% Tween-20. Blocking was performed using PBS/1% Bovine Serum Albumin (BSA) for 1 hour at 37°C. Serum samples were diluted 1:100 in dilution buffer (PBS/1% BSA/0.05% Tween-20 (PBT)), while the reference EMS sample was diluted at 1:50 and 1:200. A standard curve was generated starting at 2 IU/mL, with a 1:2 serial dilution. After incubation for 1 hour at 37°C, RF-IgM levels were detected using horseradish peroxidase (HRP)-labeled Goat-anti-Human IgM (Millipore, AP114P), diluted to 0.3 µg/mL in dilution buffer. Detection was performed by adding 2,2′-azino-bis[3-ethylbenzothiazoline-6-sulfonic acid] (ABTS) with 0.015% hydrogen peroxide (H₂O₂), and absorbance at 415 nm was measured using a Spectramax E3-46 microplate reader. The assay was monitored with measurement stopping at approximately 2 hours or when the highest standard reached an OD of 3.0.

##### ACPA

Streptavidin was diluted to 1 µg/mL in Coating Buffer (PBS/0.1% BSA) and used to coat 384-well plates (15 µL/well), followed by overnight incubation at 4°C. The following day, plates were washed three times with PBS/0.05% Tween-20, then coated with 10 µg/mL of the designated peptide (CCP4, CArgP4) for 1 hour at room temperature. Plates were again washed three times before sample incubation. Serum samples were diluted 1:50 in dilution buffer (PBS/1% BSA/0.05% Tween-20 (PBT)), and the reference EMS sample was diluted at 1:50 and 1:200. Standards were prepared with serial 1:2 dilutions, with additional Standard 4 included as a reference. Pre-diluted samples and standards (15 µL/well) were incubated for 1 hour at 37°C. After washing, IgG antibodies were detected using HRP-labeled Rabbit-anti-Human IgG (DAKO, P0214), diluted 1:5000 in dilution buffer, followed by incubation for 1 hour at 37°C. Plates were developed using ABTS with 0.015% hydrogen peroxide (H₂O₂), and absorbance at 415 nm was measured using a Spectramax microplate reader. The ELISA plates were read at multiple time points over approximately 2 hours or when the highest standard reached an OD of 3.0.

##### Anti-CarP ELISA

Anti-CarP IgG antibodies in sera from both patients and controls were detected using ELISA, following the method previously described ^[153]^. Nunc Maxisorp plates (Thermo Scientific) were coated overnight with either nonmodified fetal calf serum (FCS) or CarP-modified FCS. Between each procedural step, the plates were washed three times with Phosphate Buffered Saline (PBS) containing 0.05% Tween (Sigma, P1379). Blocking was performed using PBS with 1% Bovine Serum Albumin (BSA) for six hours at 4°C. Subsequently, serum samples diluted at 1/50 were incubated overnight at 4°C. To ensure standardisation, each plate included a positive control serum for anti-PTM antibodies to facilitate the calculation of arbitrary units. Following incubation, IgG levels were detected using horseradish peroxidase (HRP)-labeled Rabbit-anti-Human IgG (Dako, P0214). Detection was achieved by incubating the plates with 2,2′-azino-bis[3-ethylbenzothiazoline-6-sulfonic acid] (ABTS) and 0.015% hydrogen peroxide (H2O2) (A1888 and 7,722-84-1, both from Merck). The absorbance at 415 nm was measured using a Bio-Rad iMark microplate reader. Plates were developed using ABTS with 0.015% hydrogen peroxide (H₂O₂), and absorbance at 415 nm was measured using a Spectramax microplate reader.

**ANA** were detected using immunofluorescence on Hep2 slides using a commercial kit (Sprinter XL, Euroimmun) with a cut-off for positivity at dilution over 1/80.

###### Cytokine/CRP quantification

Cytokine quantification was performed using MSD chemi-luminescence assays according to manufacturer’s instruction for IL1-beta, IL-2, IL-4, IL-6, IL-8, IL-10, IL-12p70, IL-13, IFN, TNF, and IL-17A and C-reactive protein (CRP). For several cytokines (IL-1β, IL-2, IL-4, IL-8, IL-12, IL-13) results were below the limit of detection for a substantial number of cases. Lower limit of detection was reported.

#### Flow cytometry

Flow cytometry was performed on cryopreserved cells according to standard procedures. Briefly, frozen cells were thawed and resuspended in phosphate-buffered saline (PBS), then centrifuged at 600 g at room temperature for 10 minutes. Cells were resuspended in 50 µL of blocking buffer (10% human immunoglobulin and 30% mouse serum in PBS) and 50 µL of BD Horizon™ Brilliant Stain Buffer (where needed), distributed into flow cytometry tubes with various antibody cocktails across 7 panels (markers, and fluorochromes below), and stained for 30 minutes at 4°C. Cells were washed, and data were acquired using a Cytoflex-XL flow cytometer (with 7-aminoactinomycin D (7-AAD) as a viability dye). Gating strategies for each panel are presented in the respective figures.

In subsets known to correlated with age (naïve, memory Treg) data were normalised as previously reported ^[33]^ as detailed below. A subsequent batch of samples was analysed with a reduced number of markers in the T-cell panels only.

There are age relationships between naïve, memory T and B-cells and with Treg frequencies as shown below in **Figure-S1**. We established regression equations using healthy controls data to normalised frequencies, as indicated on the plots. Y represents the expected subset frequencies at defined ages. For example:

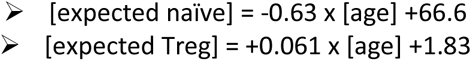

Subset frequencies reported are then normalised and reported as the difference in % cells compared to the expected value in heath.

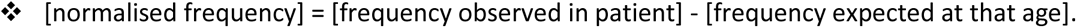

The latter being calculated from the age-subset frequency correlation described above.

Whole blood count were not available to transform data into cell number/mL blood. Data analysis comparing cell numbers versus % of cells were previously discussed ^[33]^ and did not impact conclusion or statistical results while adding data manipulation.

#### Markers included in the flow cytometry panels

|  | panel A | panel B | panel C | panel D | panel E | panel F | panel G |
| --- | --- | --- | --- | --- | --- | --- | --- |
| Fluorochrome | NK and NKT | T-cells subsets | Exhausted T-cells | B-cells | Monocytes | CYTOLITIC | Treg |
| BU395 | CD3 | CD3 | CD3 |  | CD14 | CD3 | CD3 |
| BV421/450 | KIR3DL1 (CD158e) | CCR7/CD197 | CD38 | CD38 |  |  |  |
| V500 |  | CXCR6 /CD186 |  |  |  |  |  |
| FITC | KIR2DL1 (CD158a) | CD57 | TIM3 | CD24 |  |  | FOXP3 |
| PE | CD16 | CD45RO | PD1/CD279 |  | CD16 |  |  |
| BV605 | HLA DR | HLA DR | CD44 |  | HLADR |  |  |
| CF594 |  |  | LAG3/CD223 |  |  | GranzymeB |  |
| APC |  |  |  |  | CD11c | Perforin | CD127 |
| PE-CY5 | CD56 | CD45RA |  |  | CD1c | CD56 | CD25 |
| A700 | CD62L | CD62L | CTLA4 /CD152 | CD138 |  |  |  |
| BV785 | CD27 | CD27 | CD161 | CD27 |  |  |  |
| PE-CY7 |  | CD4 | CD4 |  |  |  | CD4 |
| BV805 | CD8 | CD8 | CD8 | CD19 |  | CD8 |  |

**Figure S1:**
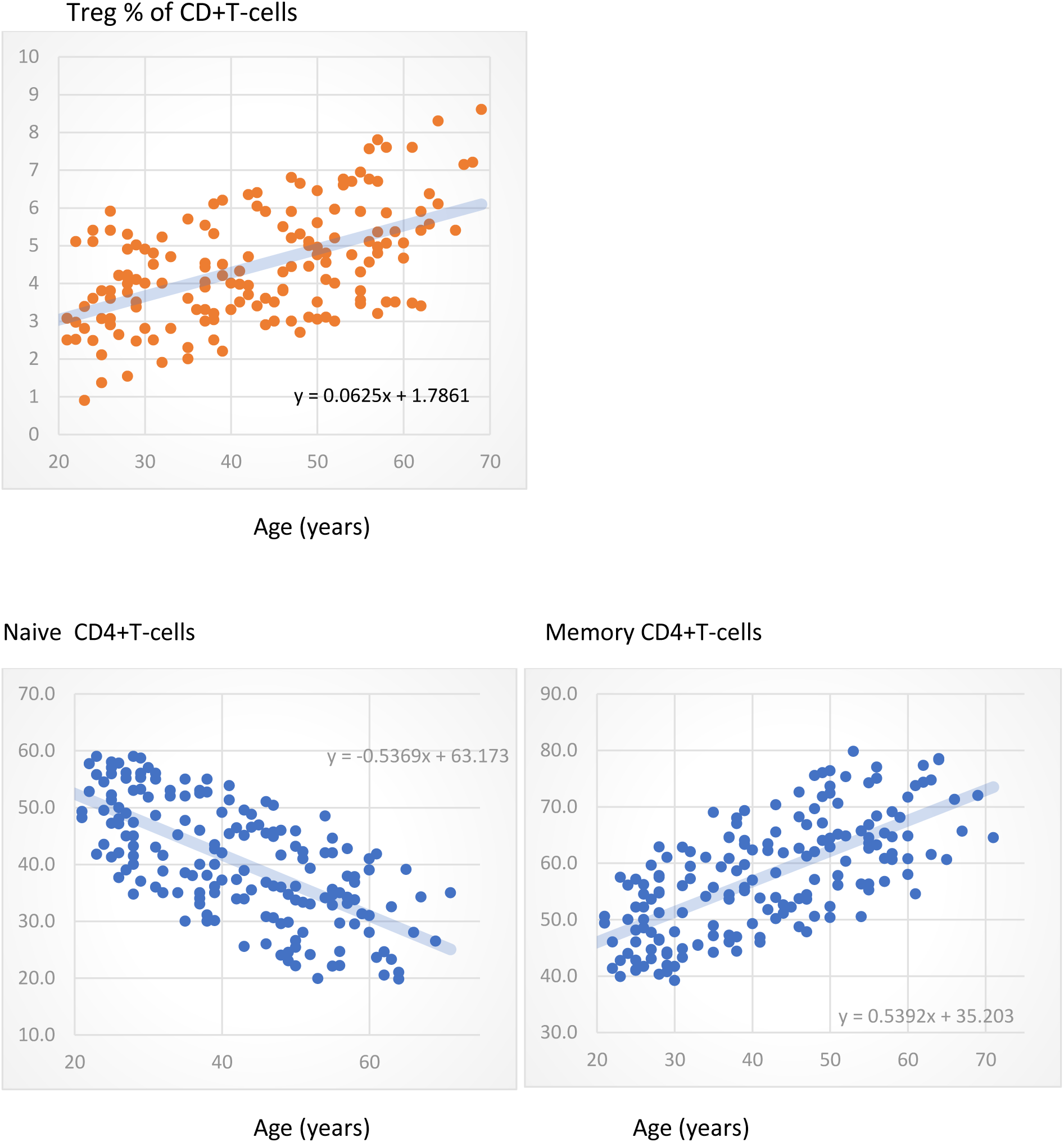
Relationships between age and % of naïve, memory and Treg subsets of CD4+Tcells.

#### Fatigue Biomarkers

We measured blood concentrations using commercial assays following manufacturer’s instructions for phosphate inorganic (Abcam, Cambridge, UK), intact FGF23 and total FGF23 (Quidel Ortho, CA, US), Lipocalin-2 (Epitope Diagnostics, CA, US).

#### microRNA RT-qPCR

Total RNAs (including small RNAs), were extracted from 200 uL of serum samples using MiRNeasy Serum/Plasma kit (Qiagen) and the QIAcube automated instrument. Prior to extraction, 5pM of cel-mir-39 were added as exogenous control for normalisation. Samples were resuspended in 15ul of RNase free water. Extractions were carried out on 2 consecutive days to minimise bias. Total RNAs were converted into cDNAs with Taqman MicroRNA Reverse Transcriptase kit (Thermo Fisher) according to manufacturer’s instructions. A pre-amplification step was performed using commercial assays for the selected mir (hsa-mir-766, hsa-mir-144, hsa-mir-21, hsa-mir-671) and 2 control mir (cel-mir-39, hsa-mir-191) using TaqMan Preamp Master mix 2X kit (Thermo Fisher). Quantification of expression was then performed, using TaqMan universal II master mix (Thermo Fisher) with primer and probes from the commercial assays and run on a Viia 7^TM^ Real-Time PCR instrument (Thermo Fisher). RT-qPCR raw data were analysed using Quant Studio software and normalised to cel-mir-39 and hsa-mir-191 expressions.

The National Office of Statistics in the UK ran a survey of symptoms and characteristics of people with self-reported long-COVID over 2 years, between April 2021 and March 2023. Data are publicly available under the format of CVS files with estimates (95% CI) of % or numbers of cases on a monthly basis. We used data from 24 monthly downloads to generate the graphics in Figure-S1. This data has been published ^[151, 152]^ and our group of LC patients align well with the general UK population of LC.The data related to work status **after** Long-COVID (pie chart), were collated from response in the YC19-PRO/PROM proposing different categories: unable to work, took early retirement, student (excluding children at school), not working (no change from pre-pandemic) or was able to return to work with/without adapted conditions. This is in agreement with the data on work-status by the National Office of Statistics published ^[151, 152]^ which suggested similar representation of patients able to work, studying, retired or not employed (∼15%) and a higher percentage of “inactive” patients (∼40%).

**Figure S2.**
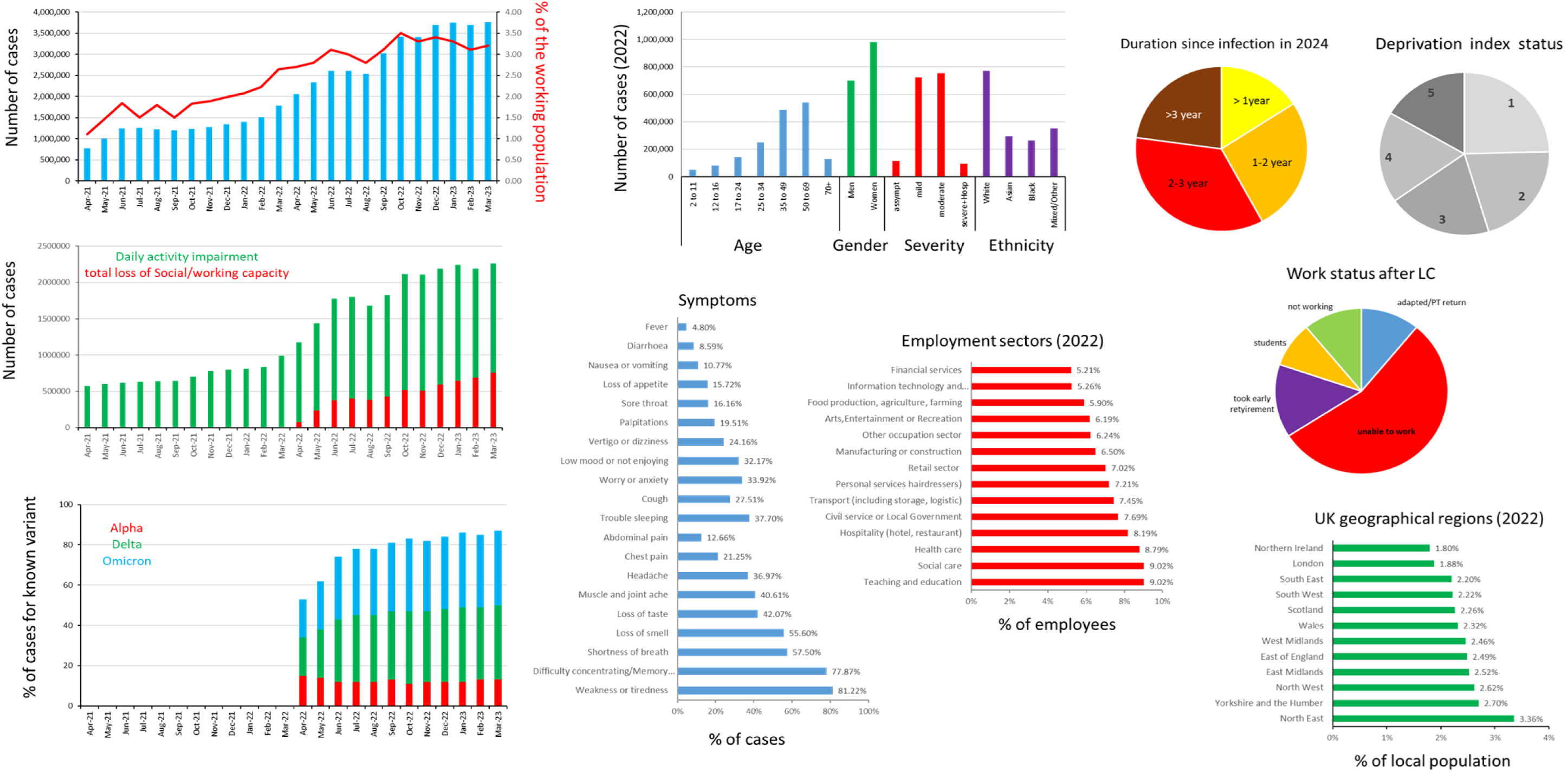
Data gathered from monthly reports by the UK Office of national statistics on self-referred cases of Long-COVID (2021-23).

**Figure S3:**
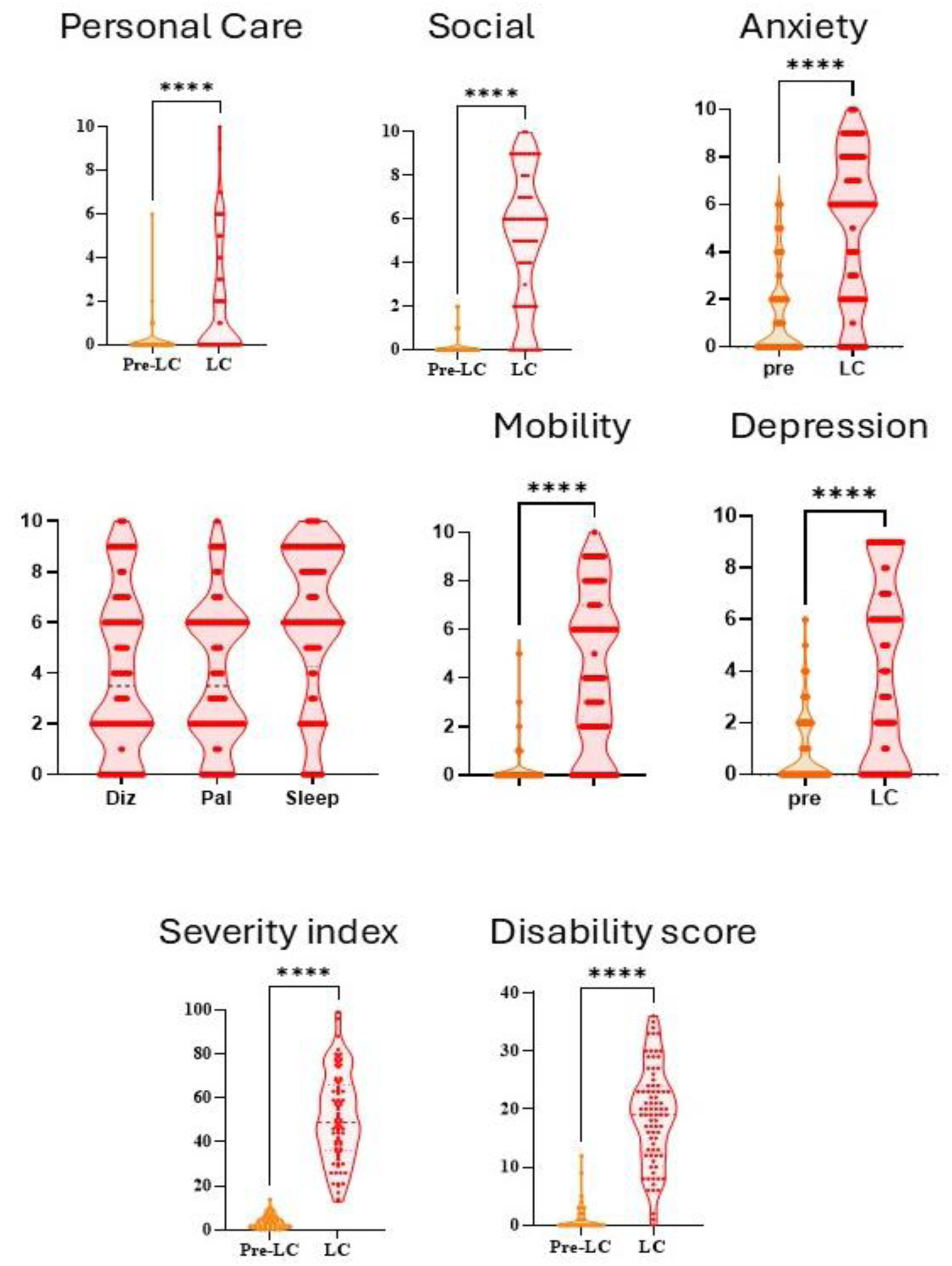
additional PRO reported via the YC19-questionnaire in LC patients, pre-pandemic (orange) and during LC (red). PRO are scored 0 => 10 where 10 is worst. Only 9/82 (11%) reported an absence of cognitive issues, while 8/82 (9.7%) indicated a pre-pandemic history of mental well-being, exacerbated by LC. Breathing difficulty (score >1) was reported pre-pandemic primarily during stair climbing (17%), whereas breathing at rest (51%), during activities (60%), or climbing stairs (88%) were major concerns in LC. Body-wide pain levels exhibited a wide distribution, with only 4 patients reporting mild pain pre-pandemic. Pain was localised in joints (83%) muscles (77%), or as headaches (76%). Many patients also experienced dizziness, palpitations and sleep disturbances. Physical impairment (mobility/walking) was present pre-pandemic in only 3 patients but was a significant issue in LC (81%), as was difficulty performing daily activities (97%). Personal care was less affected, with only 17% of patients reporting difficulties (low median score=2). Social life quality was markedly impaired (81%).

#### Chronic infection

Several hypotheses have been proposed to explain LC, one frequently cited suggesting the persistent presence of live viral particles responsible for the low-grade inflammation, rather than a "sterile" disease characterised by complete viral clearance. Viral shedding, evidenced by the presence of SARS-CoV-2 RNA in stool samples, was observed in 95% of hospitalized cases ^[154]^, lasting at least 4 weeks after infection in patients who subsequently fully recovered, up to 12 weeks in 13% of participants, but only in 4% cases after 7 months (associated with lingering nausea) ^[155]^.

Here, we employed a surrogate measure for viral persistence by detecting the nucleocapsid (N) protein, a component of SARS-CoV-2 in the blood.

**Supp-Figure-S4:**
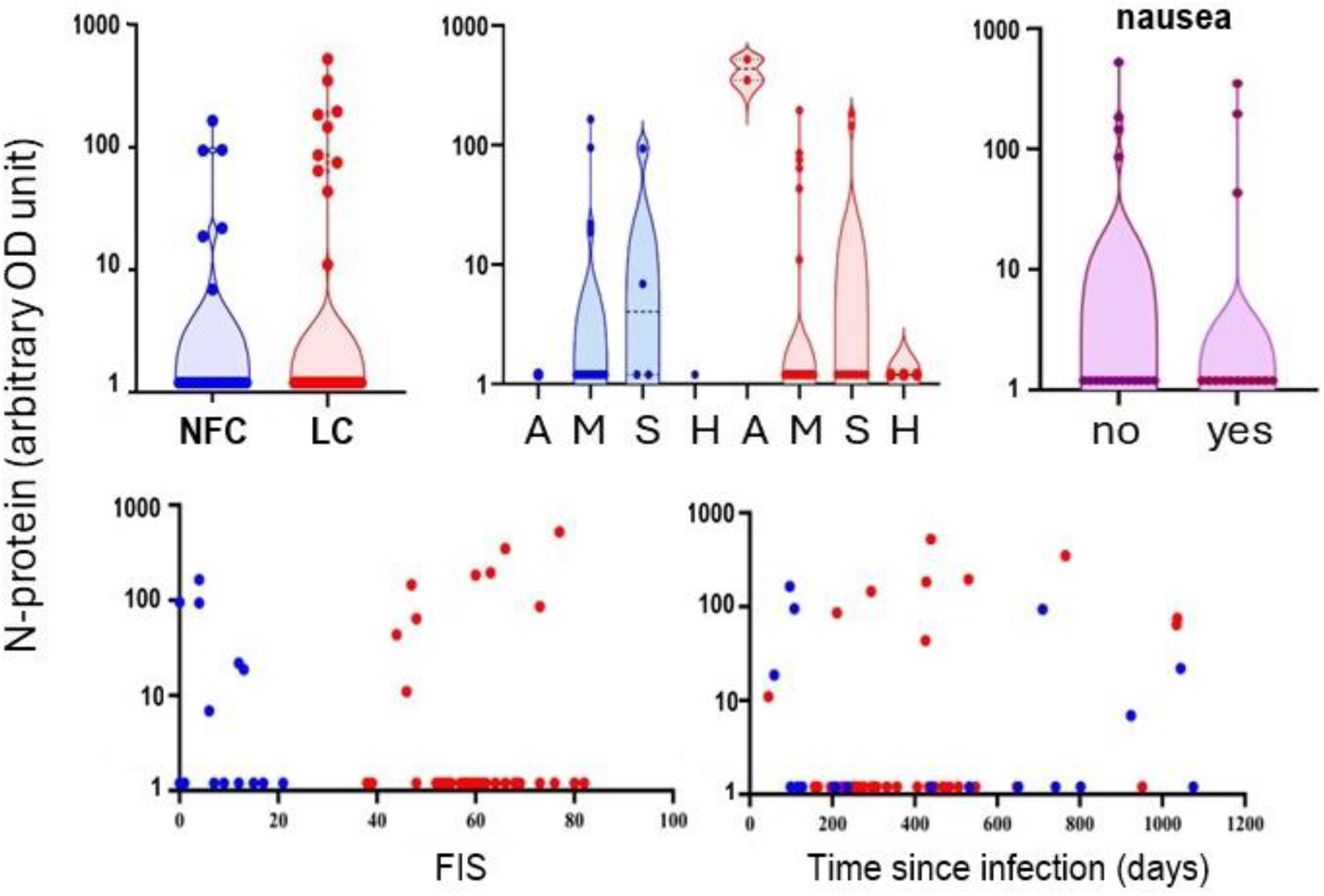
N-protein detection serum samples

In the LC cohort (**Supp-Figure S4**), the N-protein was detected in 10/39 cases (26%), with a mean optical density (OD) unit of 2.45 (arbitrary units). It was also detected in 6 out of 21 NFC (28%, median 0.95 OD). The presence of the N-protein did not correlate with the severity of the initial infection (asymptomatic, mild/moderate severe, hospitalised), FIS or time since infection. Furthermore, no association was found between N-protein detection and any specific symptoms reported in the YC19-PROM questionnaire (notably abdominal pain/nausea).

The presence of N-protein only provides information about one of the virus components and does not indicate whether fully replicating viral particles are still present. Nonetheless, despite small numbers, our data argue for a non-specific occurrence.

Given that we observed elevated levels of both IL-1β and IL-18 in LC, we investigated the potential activation of the NOD-like receptor family pyrin domain-containing 3 (NLRP3) inflammasome, which mediates caspase-1 activation and the subsequent secretion of these cytokines. To this end, we measured specks levels of apoptosis-associated speck-like protein containing a CARD (ASC), a marker of NLRP3 inflammasome activation.

**Supp-Figure S5:**
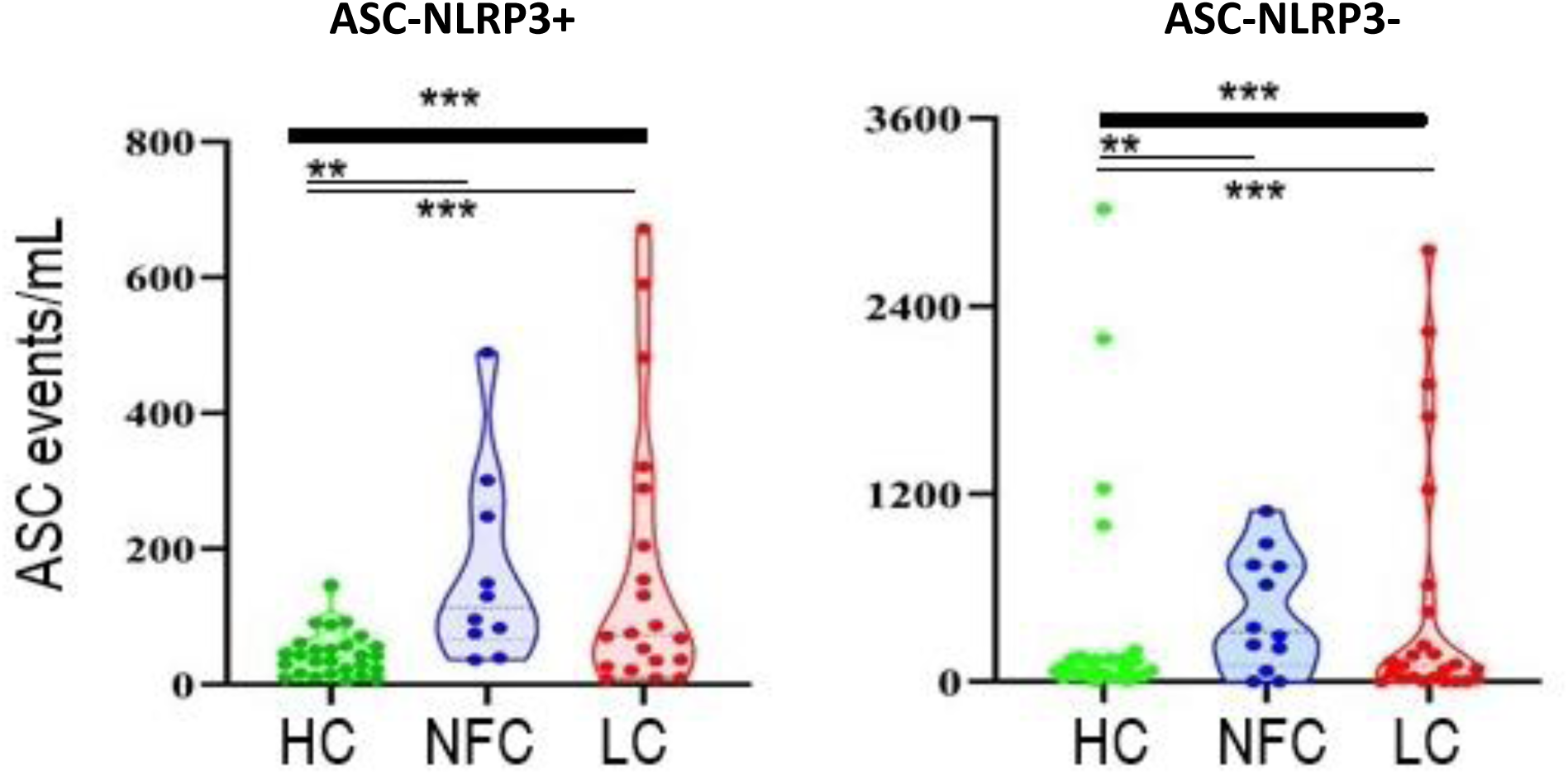
Apoptosis-associated speck-like protein containing a CARD (ASC)

ASC were quantified in serum samples from 23 LCP (red) and 12 NFC *blue) and 30 pre-pandemic HC (green). We detected increased levels of NLRP3-positive ASC specks in both NFC and LC compared to pre-pandemic (p<0.0001), but not between them. Similar results were observed in the levels of NLRP3-negative ASC specks.

##### Lymphocytes subsets

We investigated multiple lymphocyte subsets suing flow cytometry. Due to constraints with amount of material/funding, we performed a pilot lymphocyte phenotyping on 16 LC and 11 NFC, using frozen PBMCs. Altogether, large differences appeared in the frequencies of the various populations analysed (detailed gating strategies), however mostly observed both in controls and patients.

###### Lineages

CD3+CD4+ and CD3+CD8+ T-cells, CD19+ B-cell, CD56+ NK-cells, CD3+CD56+ and CD14+ monocytes were analysed and showed no significant alteration between groups (as % of circulating PBMC).

**Supp-Figure S6.**
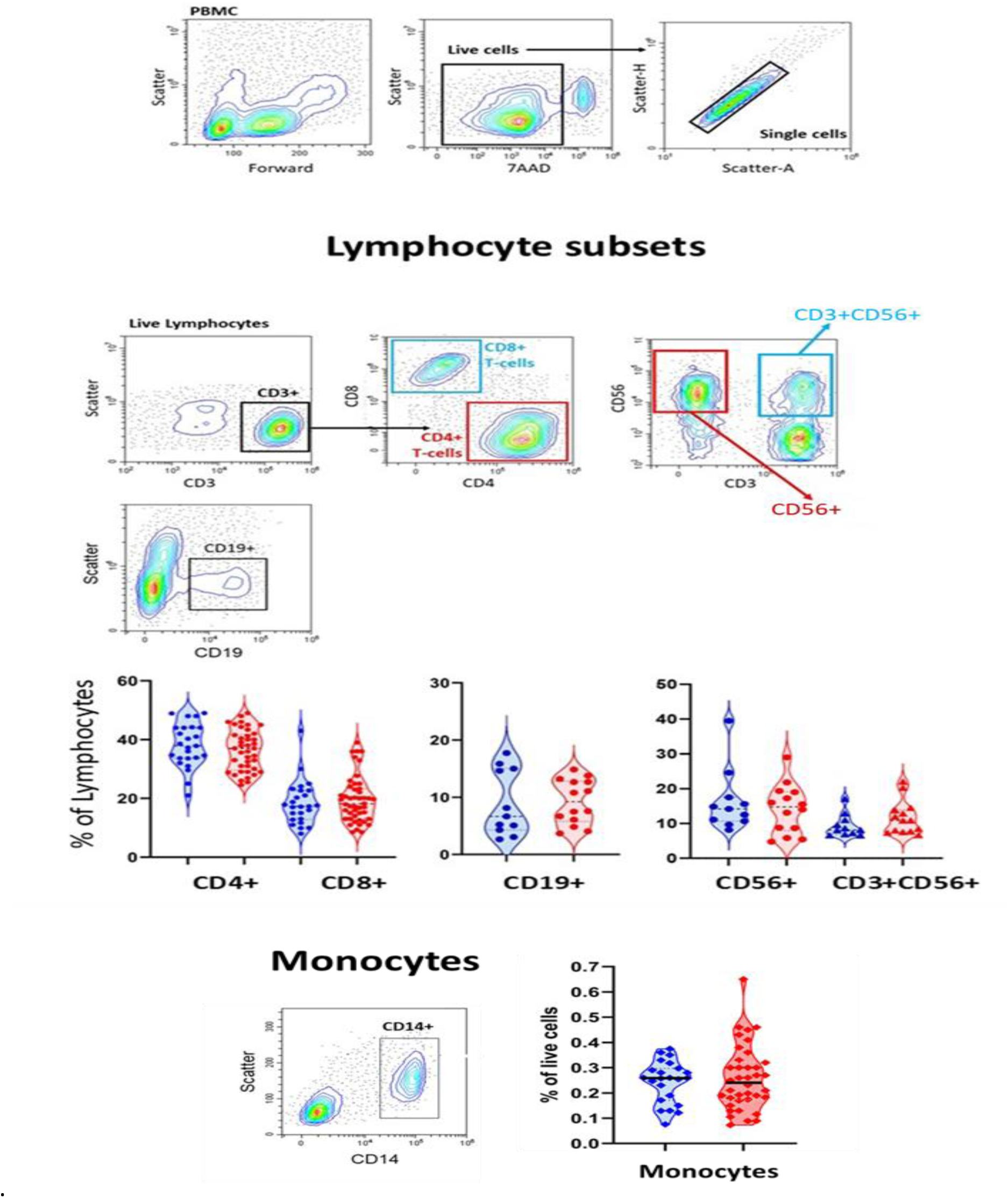
-multiple panels: Flow cytometry panel and data.

###### B-cells

No significant difference were observed for putative T2/Breg cells (CD24+/CD38^high^) and pre-plasma blasts (CD24-/CD38^high^), although a shift in the balance of naïve (CD27-) and memory (CD27+) B-cells could present.

###### Monocytes

There was no difference in CD16+ non-classical monocyte frequencies or in the levels of expression of CD11c or HLA-DR on monocytes.

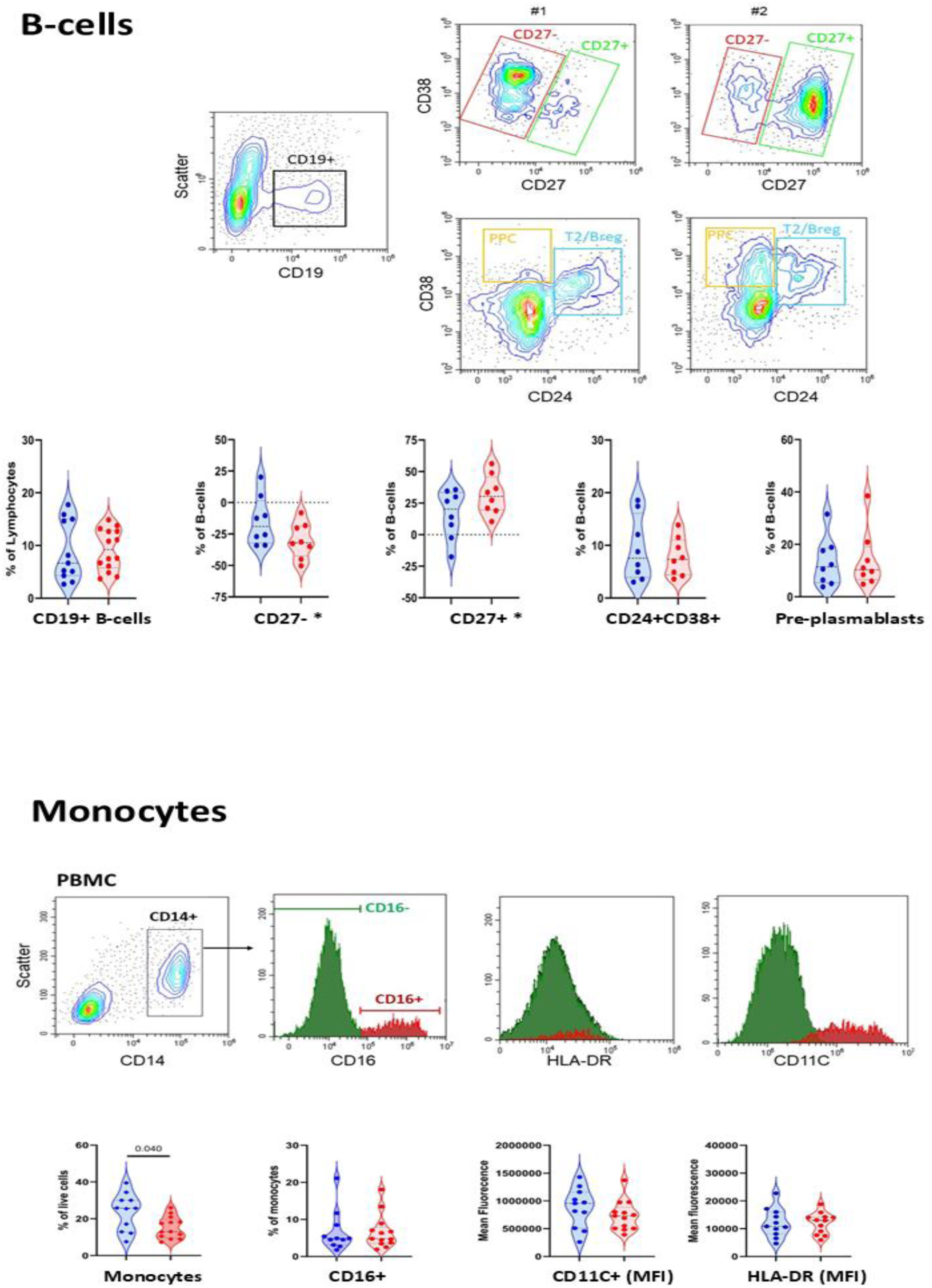

###### Cytotoxic cells

Despite large heterogeneity, there was also no difference in the capacity for cytolysis of NK, CD56+ T-cells or CD8+T-cells measured by the levels of expression of perforin and granzyme.

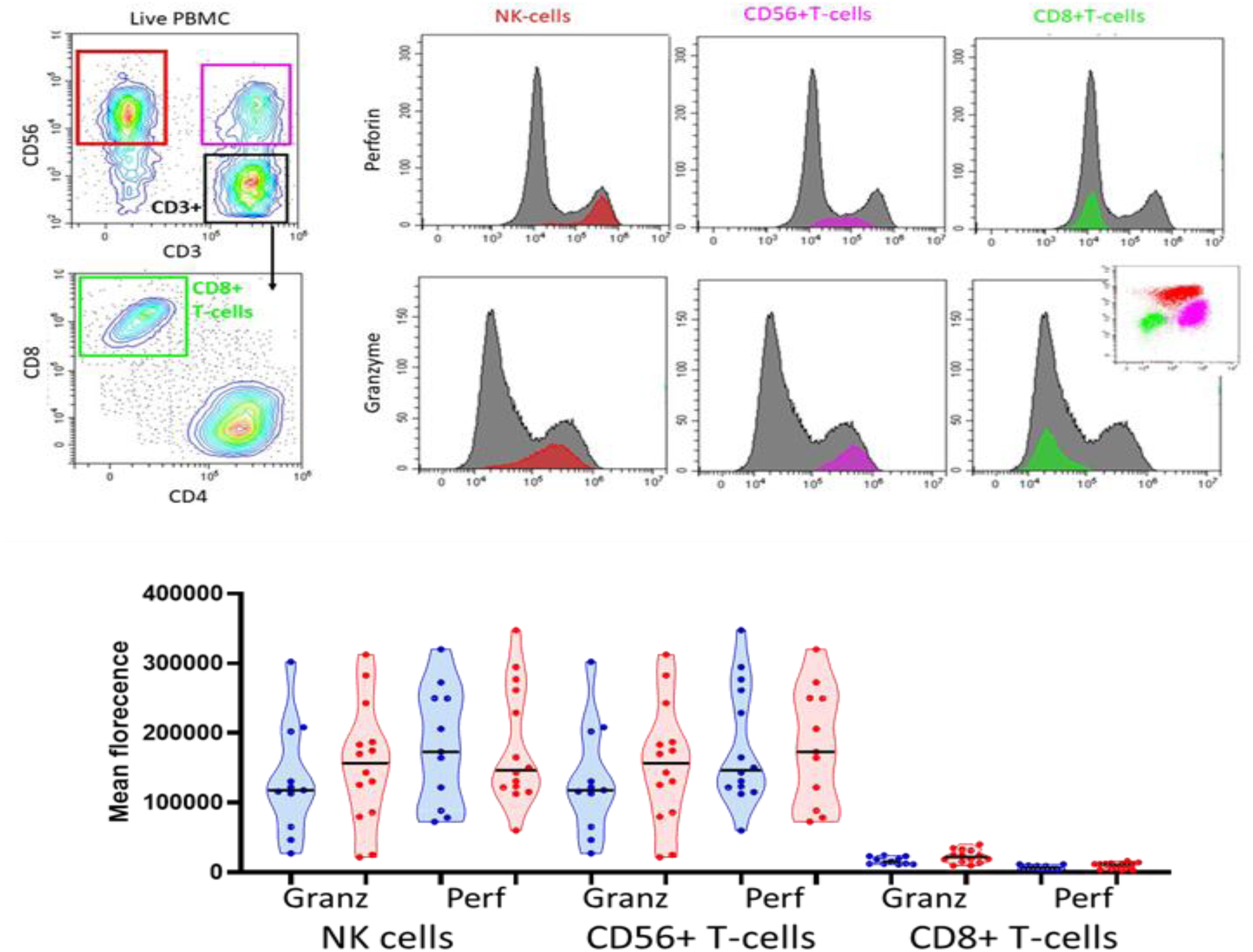

###### NK-cells

NK and CD56+ T-cells were analysed for the expression of several markers. Patterns were very heterogenous showing distinct subpopulation of cells based on KIR2DL1 and/or KIR3DL1 expression but with no difference between patients and controls. The expression of other markers (CD27, CD62L, CD8 and HLA-DR) was also heterogeneous. NK cells in LC showed significant lower CD16+ (unadjusted MWU, p=0.042) and higher CD62L+ (p=0.0007) and CD56+T-cells, lower CD27+ (p=0.0075) and CD62L+ (p=0.0011)

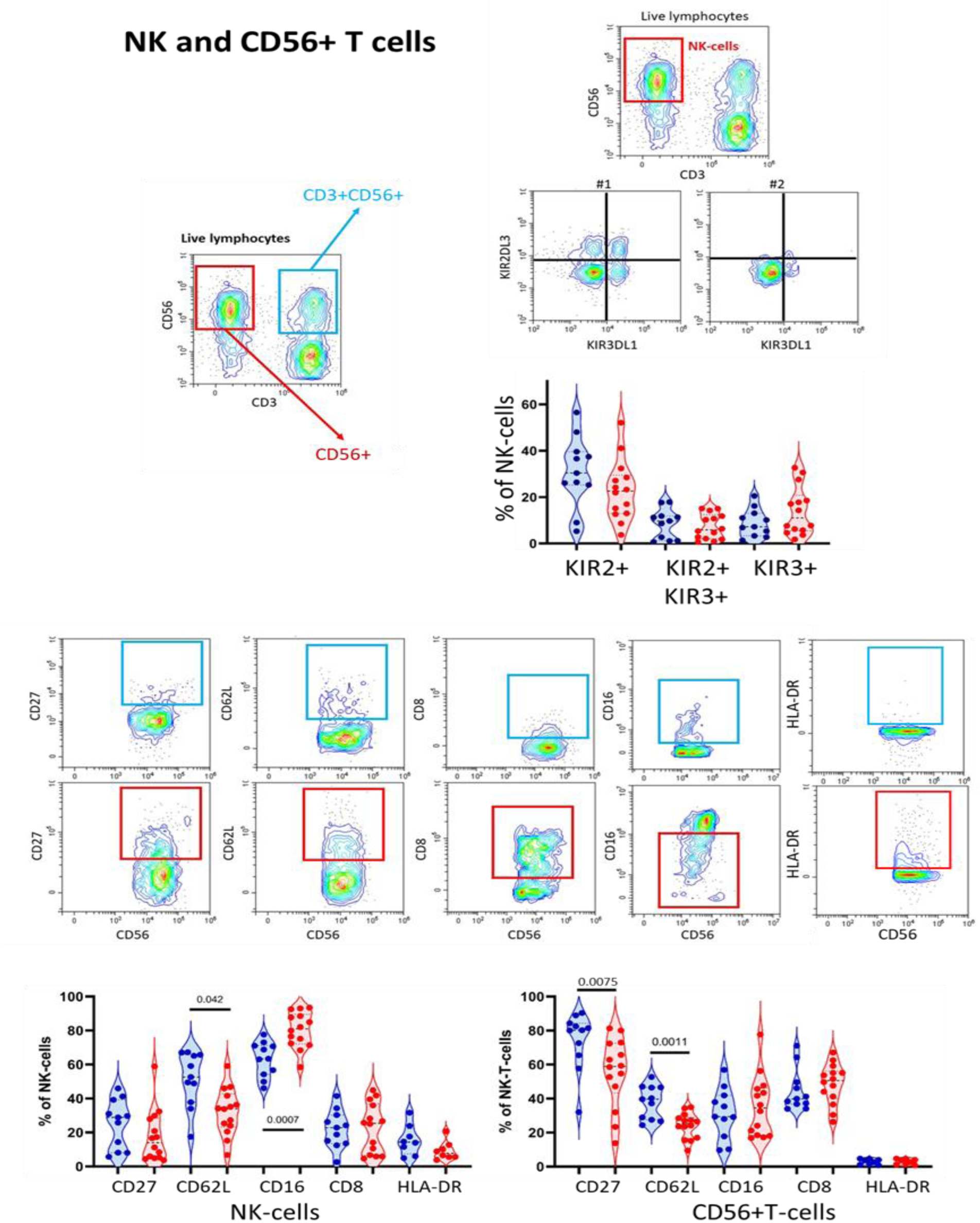

**Figure S7:**
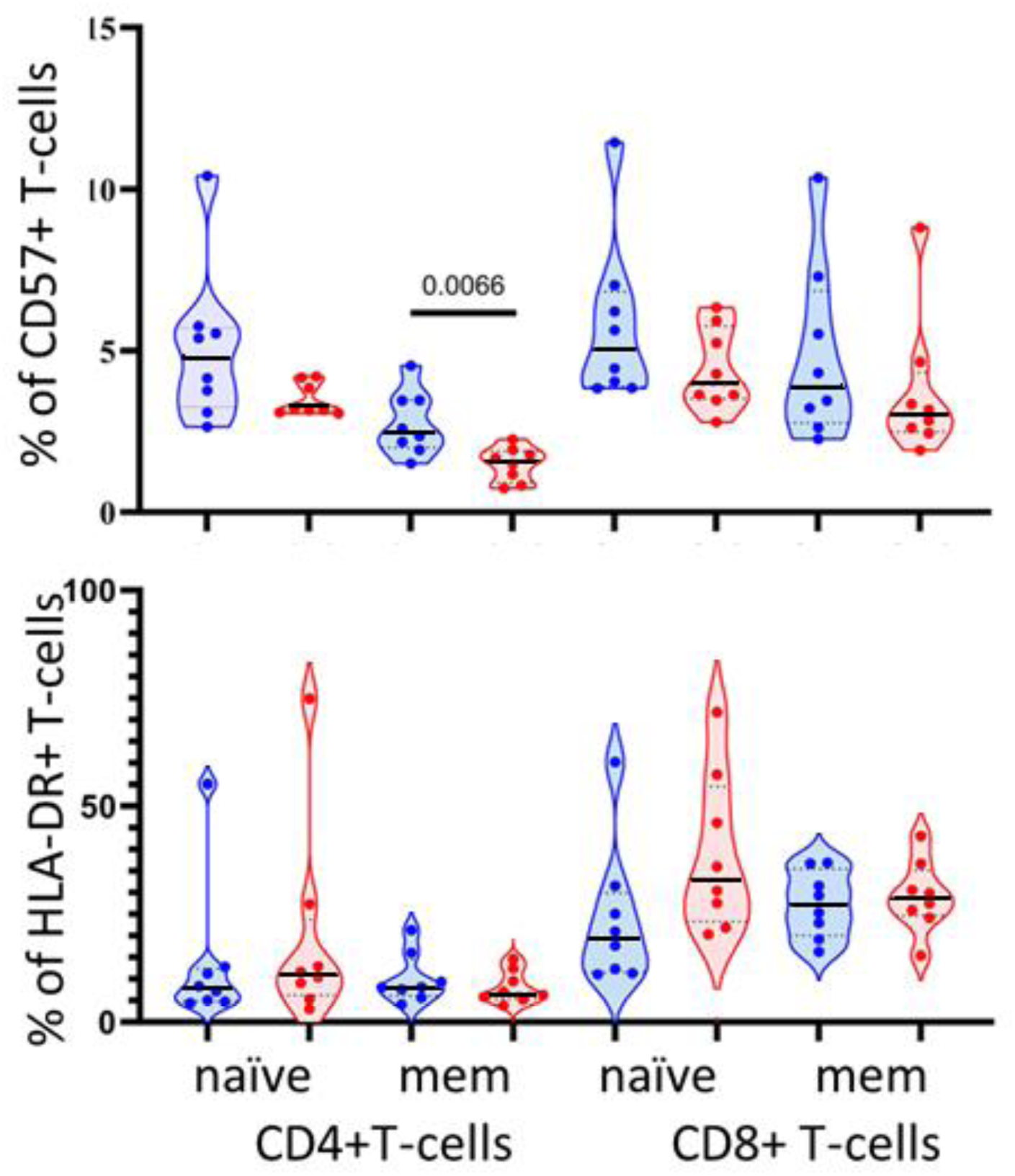
Additional marker on CD4+T-cells. Expression of various markers CD57 and HLA-DR was investigated on naïve and memory cells. Significant reduced frequencies of CD57+ memory CD4+T-cells were seen in LC (p=0.0066) and similar trends were consistently seen for all other subsets. HLA-DR+ frequencies were not significantly altered, although a higher median was seen for HLA-DR+ naive CD8+T-cells in LC.

**Supp-Table S1:**
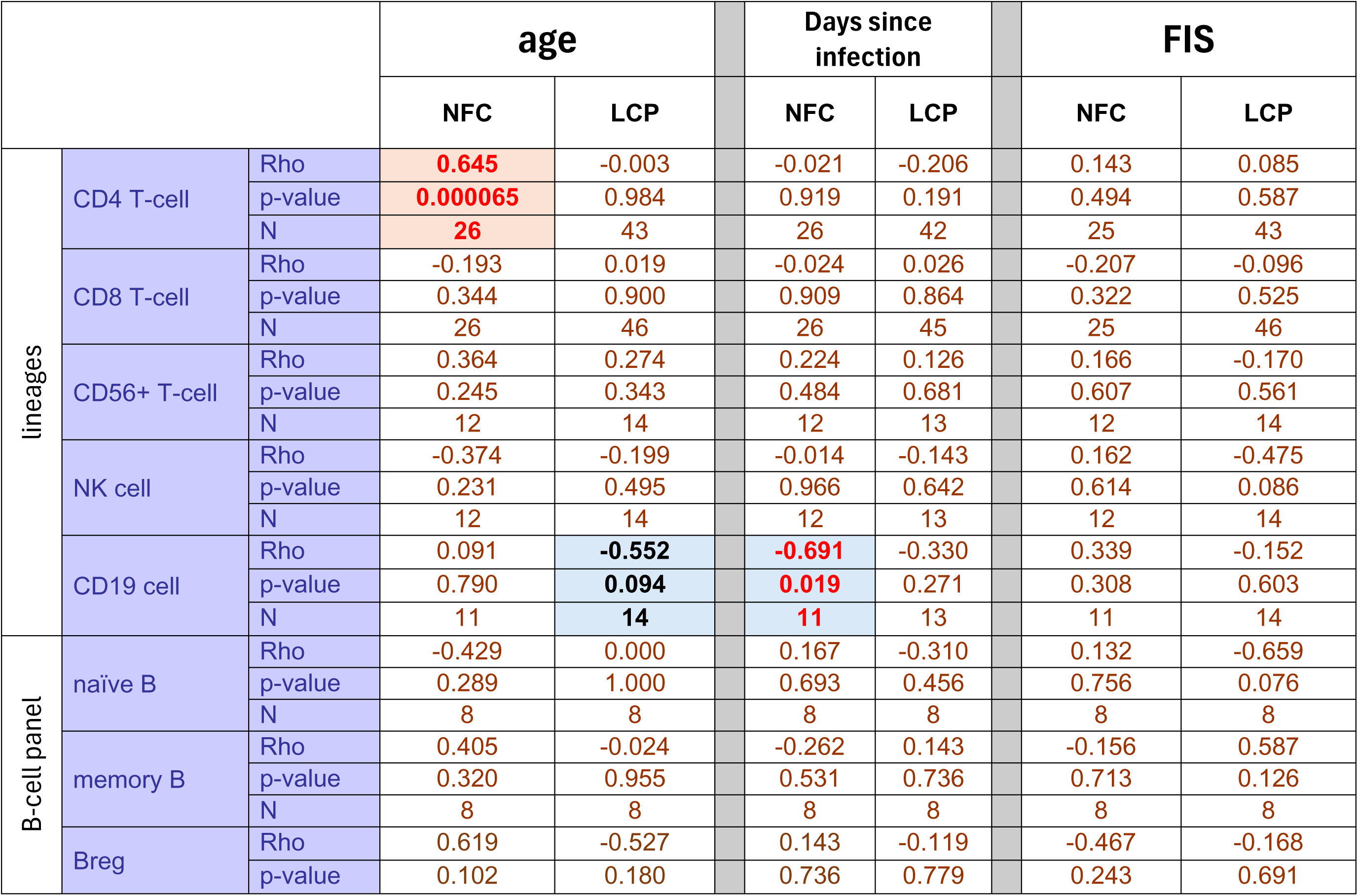
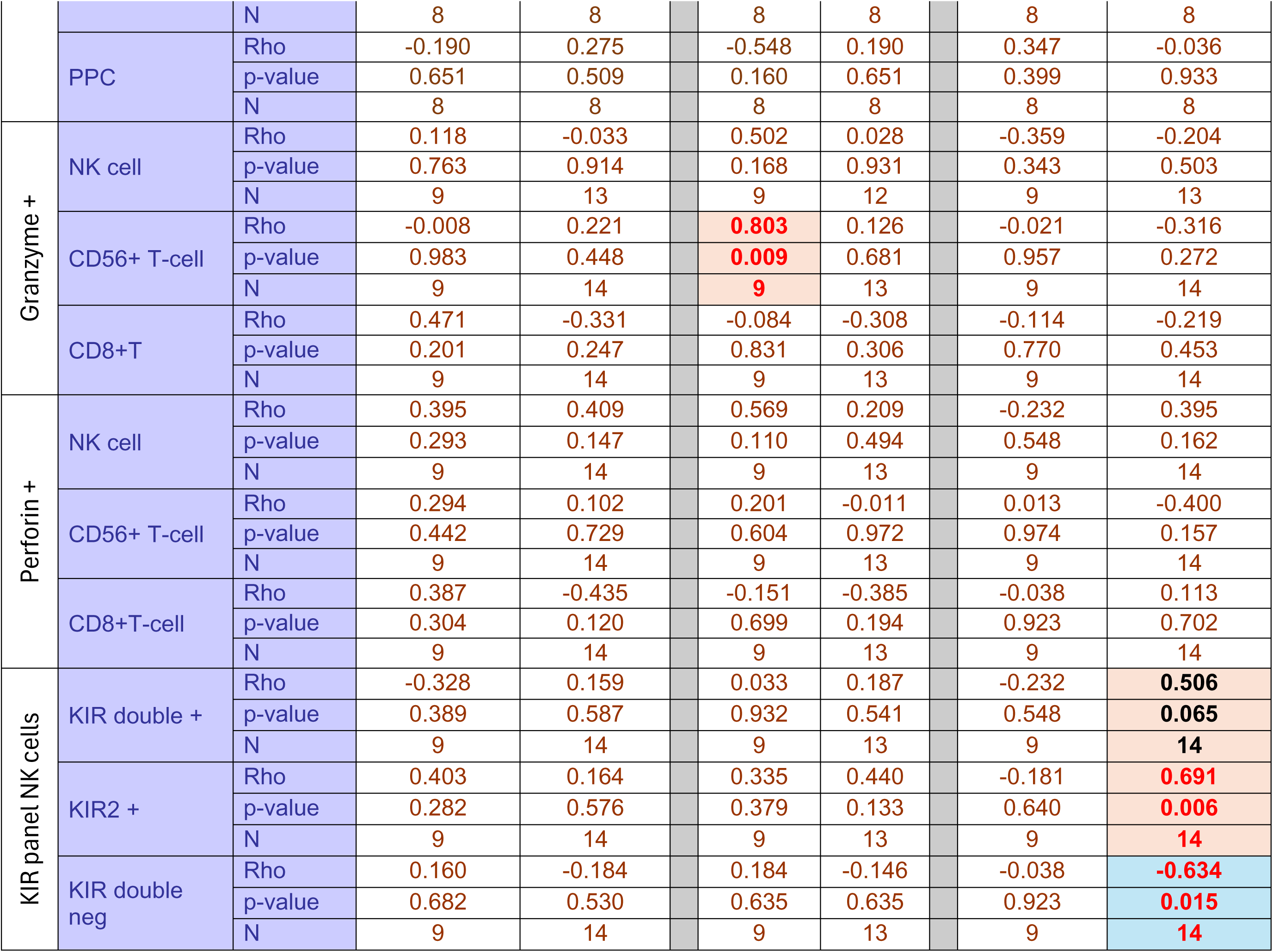
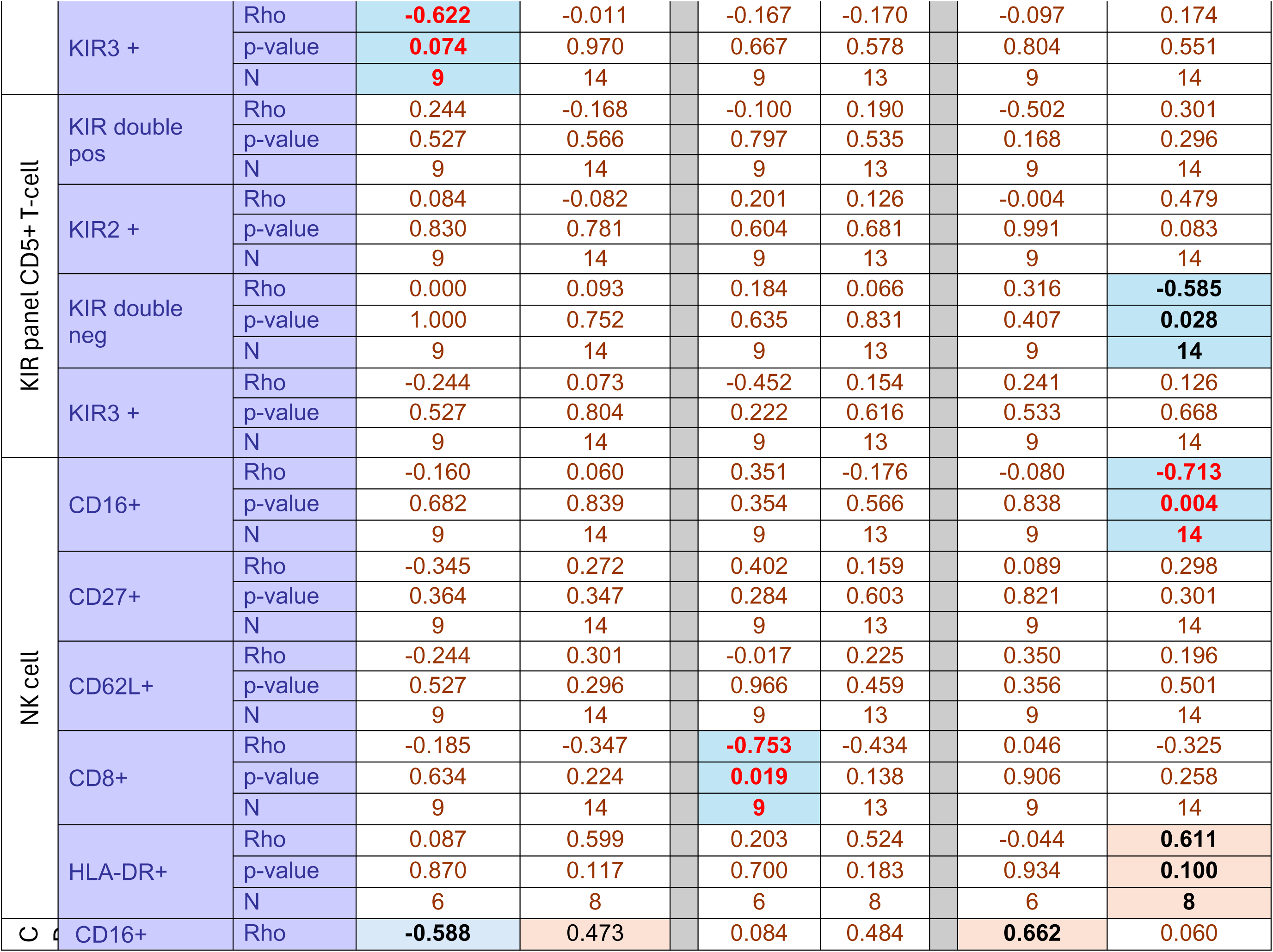
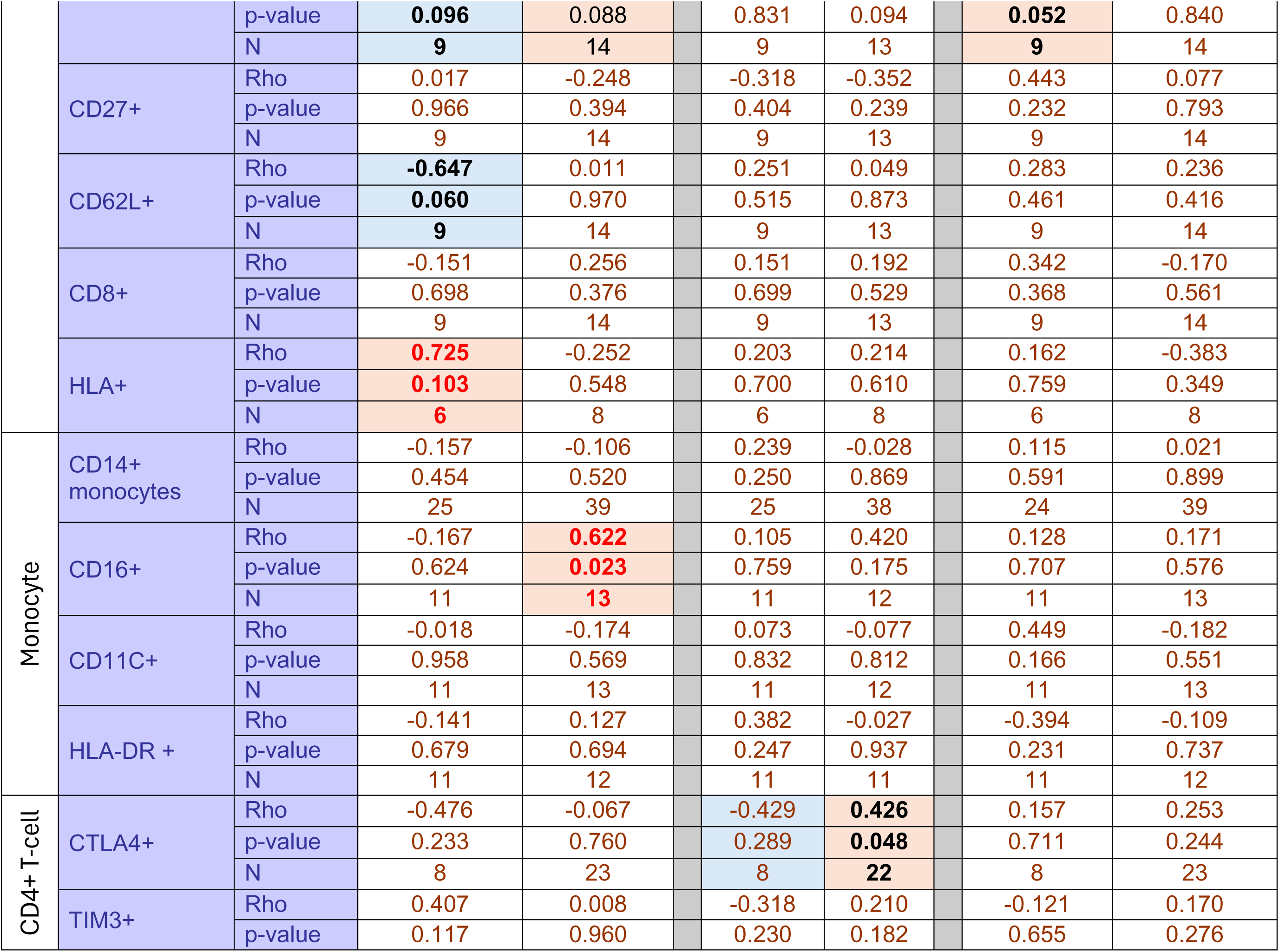
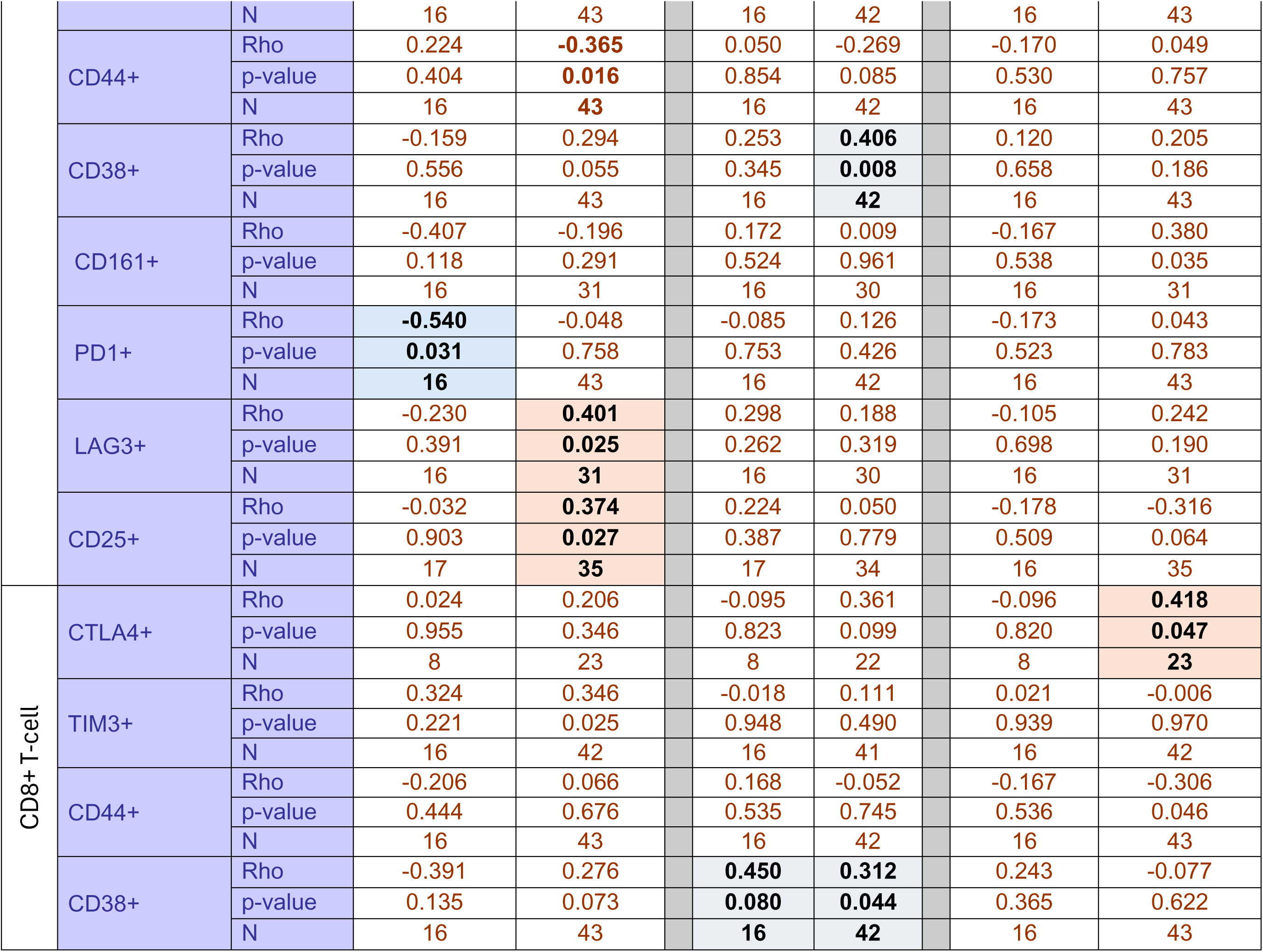
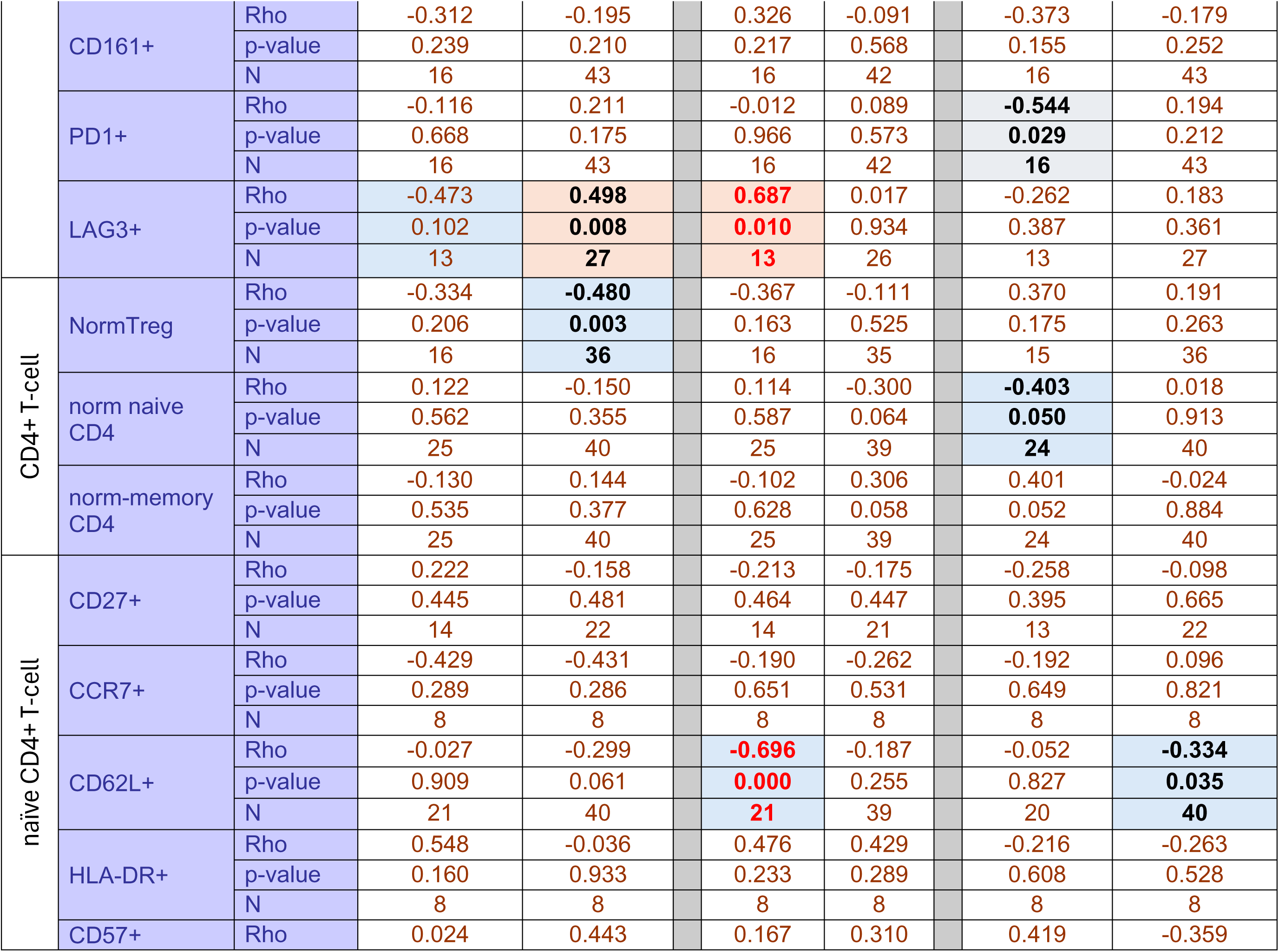
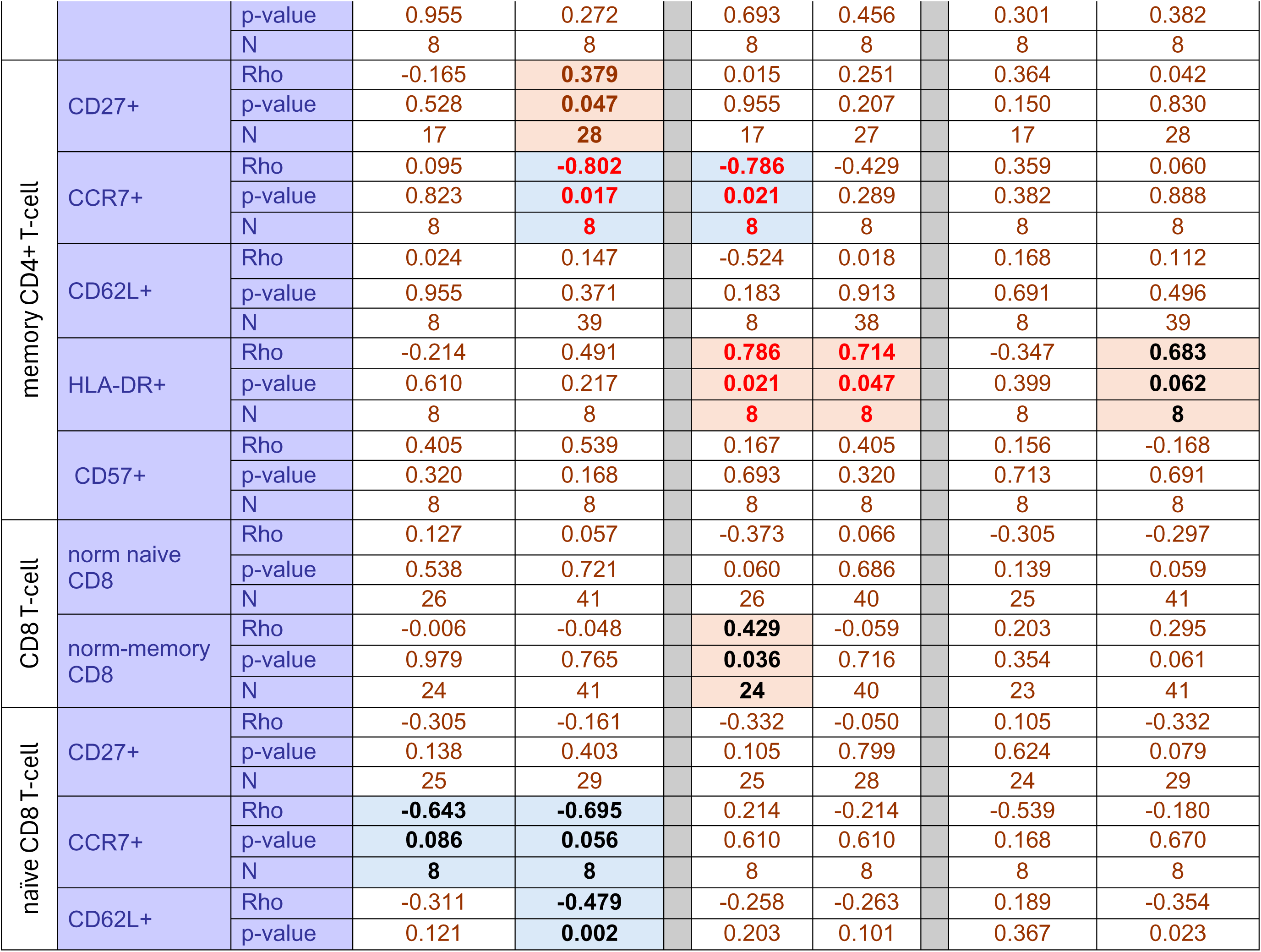
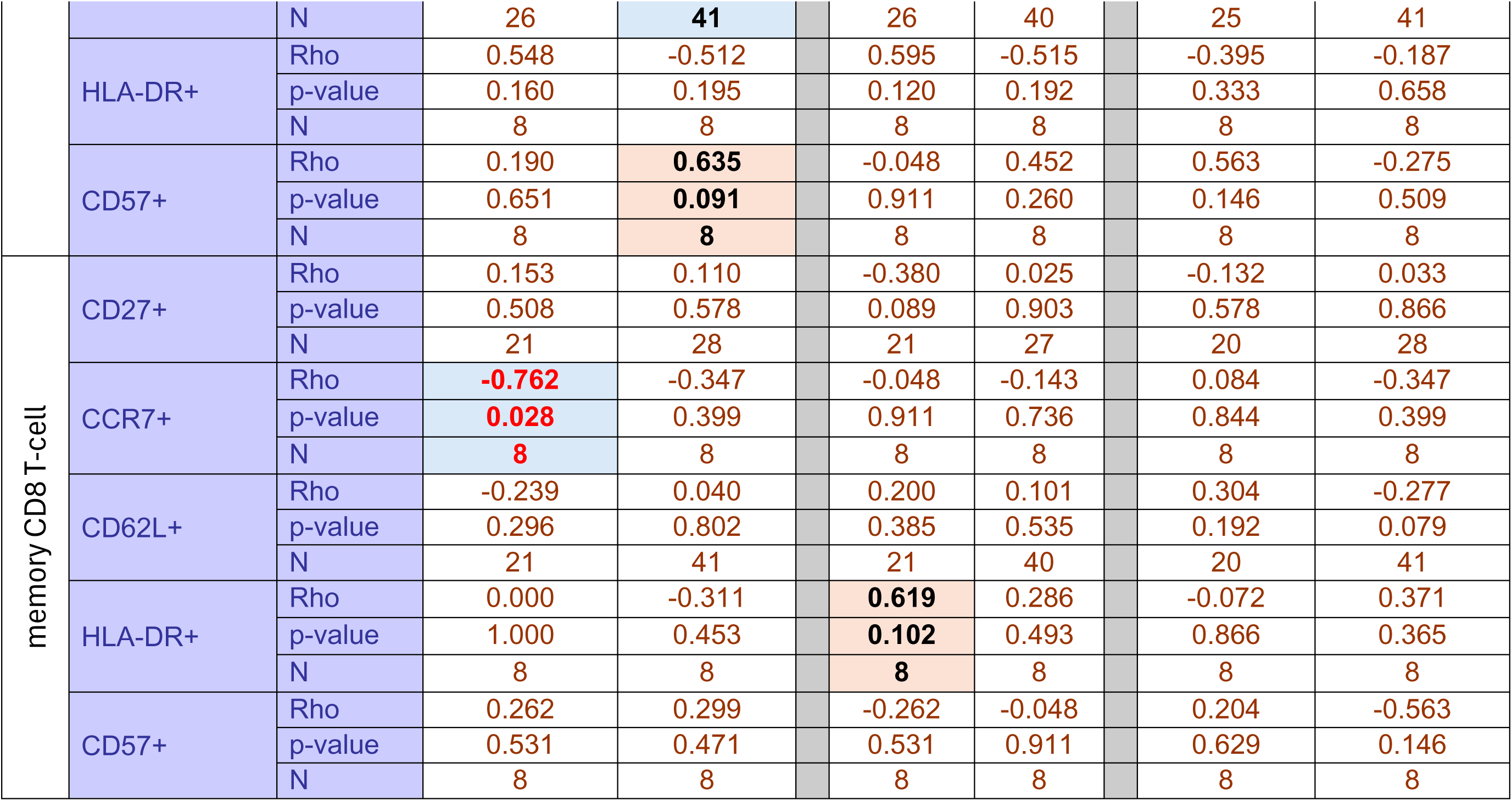
Associations between immune cell subsets and age, FIS or time since infection. Numbers being small (n ranging from 8 to 43), most association are not significant and no correction for multiple testing was made. Rho values are considered significant when above/below 0.600/-0.600 (**bold red**), and trends with rho −0.400/+0.400, when <u>p<0.100</u> (**bold/black)**. Increasing and decreasing associations are indicated in pink/blue boxes. Of note, some association tended into the same direction, others in opposite direction between controls and patients.

### miRNAs selection

We searched the biomarker literature for miR associated with fatigue, T-cell exhaustion, inflammation, autoimmunity, mitochondrial activity or neurological dysfunctions to select candidates **limiting to data in serum**. A non-exhaustive list was built with >120 candidates ^[81, 156–166]^. Overlap was observed for many candidates, 2 being associated with all 6 functionalities (miR-21, miR-144), while >50 miRs were common to at least 2 of them. We selected 4 miRs (miR-21, miR-144, miR-671, miR-766) to test based on being expressed by immune cells.

**Supp-Figure S8:**
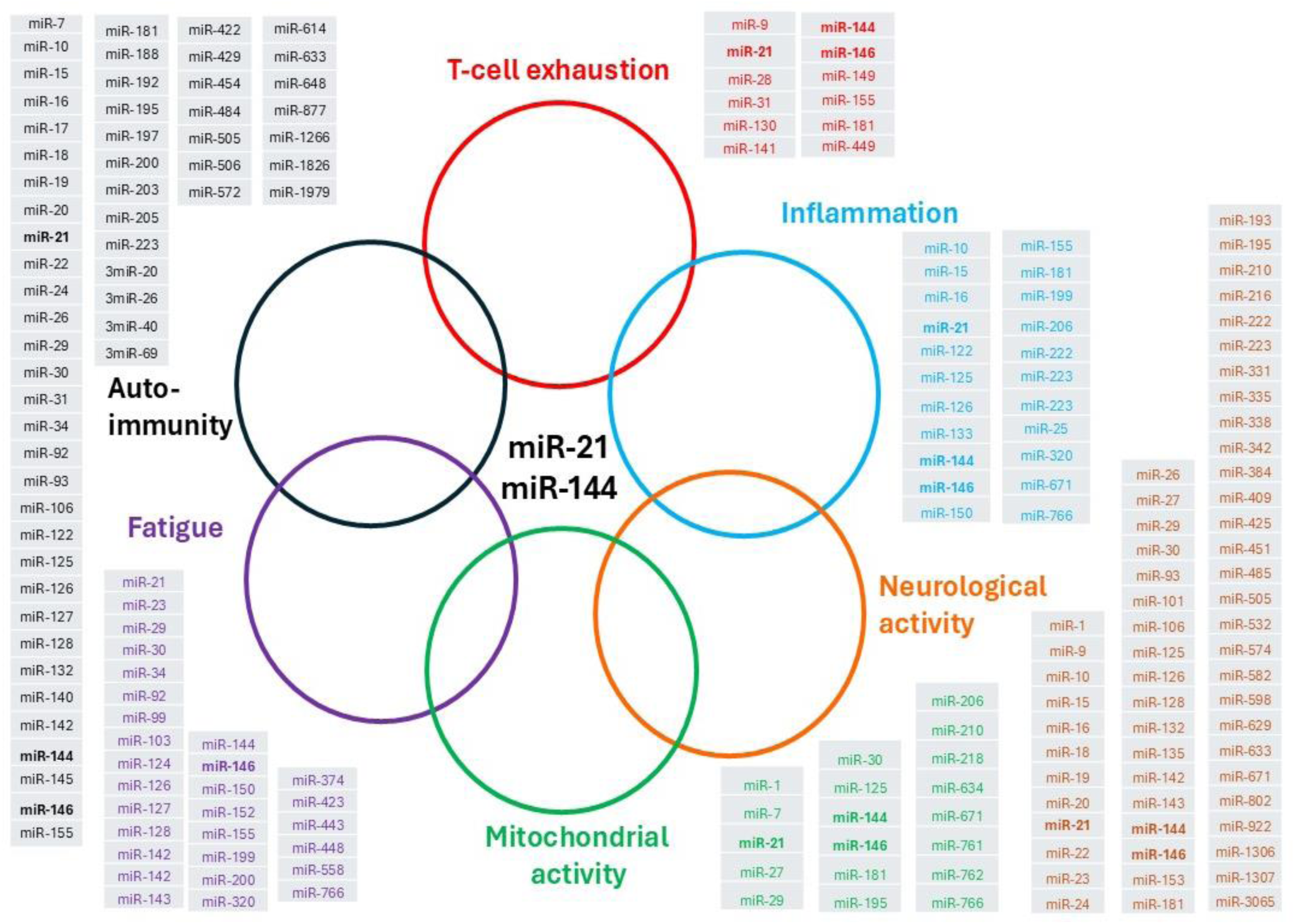
Overlap in miRNAs reported to be involved in various physiological pathways.

**Supp-Figure-S9:**
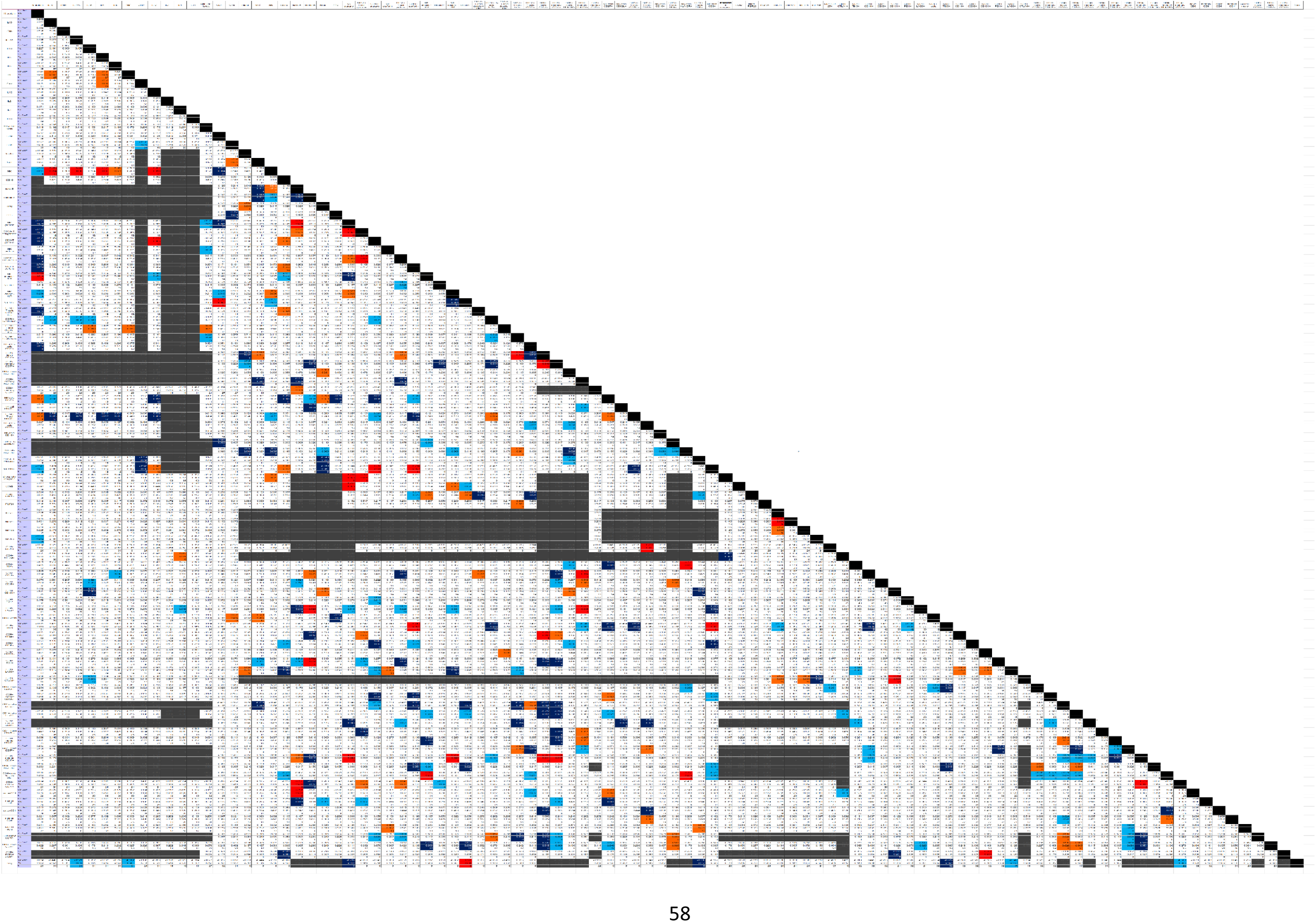
Correlation Matrix between candidate biomarker. (see supplementary excel for details of the numbers/legends). Red box rho>0.600, orange rho>0.500, light blue rho<0.500, dark blue rho<0.600. grey n<8. Blacked-box indicate missing correlation deleted due to small numbers (n<8).

**Figure S10:**
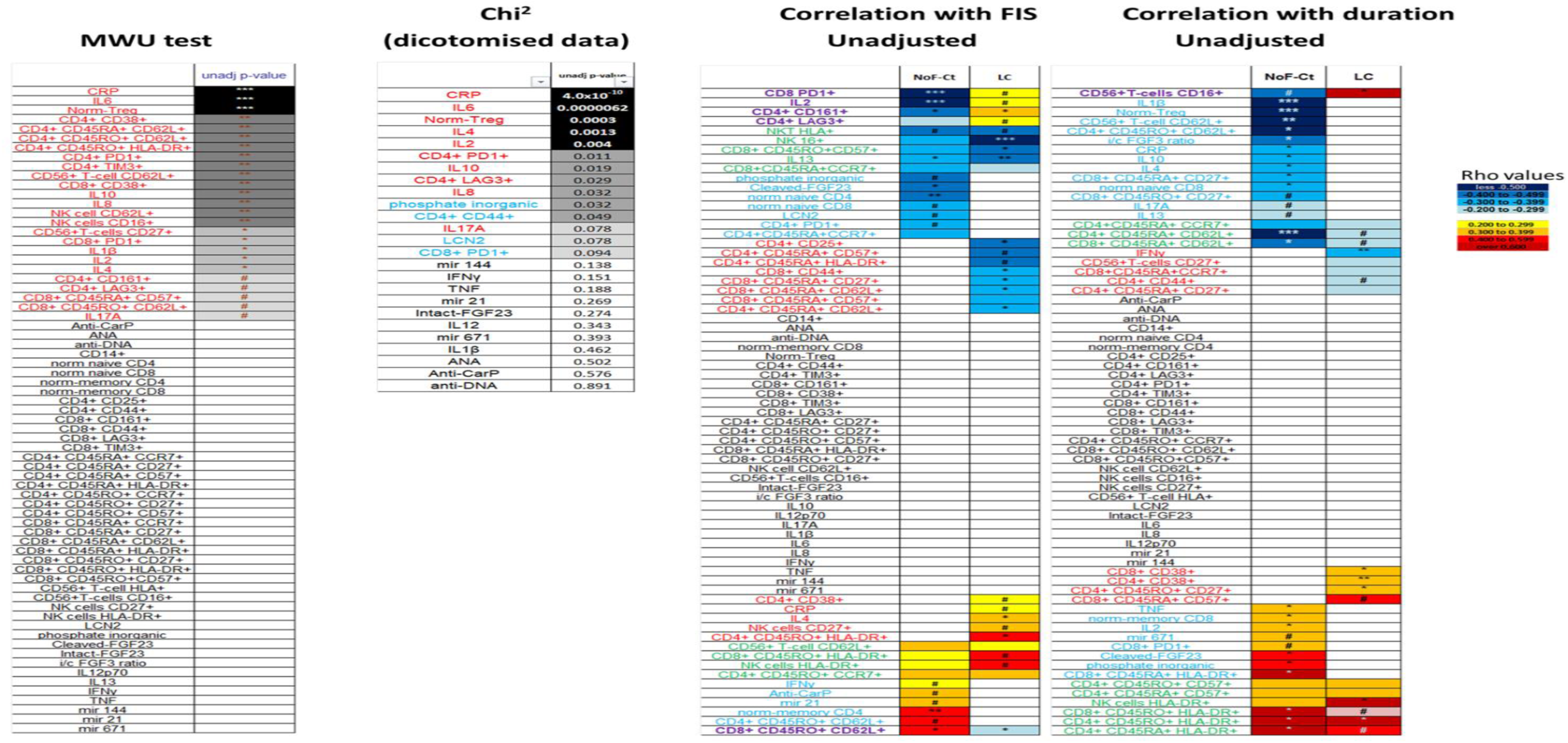
Selection of potential biomarkers. In the pilot phase, technical platform showing potential difference for some candidates between groups were included (MWU, n=25 controls/patients, left panel). Some technologies (in particular Flow cytometry) generated data for biomarkers that were not selected but part of the platform. More samples tested in the 2^nd^ stage, but not all candidates could be pursued due to limitations imposed by availability of material/funding, resulting in variable n (test results are presented combining phase 1+2 samples). **Left panel** - Comparing controls and patients, 20 biomarkers were significant (15 with p<0.001), and 5 more showing trends (). **Middle panel** - Dichotomisation of data for 25 biomarkers with a skewed distribution allowed to perform Chi2 tests, selecting the same biomarkers, with an additional 4 to the 1^st^ selection. **Right panels** - Association with FIS and duration since infection (days) are presented in the right panels, with correlation coefficient Rho colour coded. P-values for MWU or correlation: *** p<0.0001, ** p<0.001, * p<0.05, # p< 0.100. Biomarkers with opposing relationships between Ct/LC are indicated in **purple**, those with similar ones in green, some only present in controls in blue or in patients in red. Note: biomarkers with no significance are ordered alphabetically in the left/right panels.

#### Biomarker of Long-COVID

Many cases had missing data due to shortage of serum/funding and PBMC not collected in 25% of LCP; hence data were **not** missing at random (confirmed with a Little’s MCAR test). We therefore decided to use only the 28 variables selected above (by MWU or Chi^2^) for a binary logistic regression to predict LC. Data imputation was carried out using the multiple imputations function in SPSS only for these 28 variables. Pooled analysis was performed, and OR were not affected by the imputation process. Binary logistic regression was performed, using a forward method to select the best predictors. We proceeded to analysis in Serum biomarkers (n=127), cell subsets (n=69) and then combining the selected biomarkers (n=59).

**Table S2:**
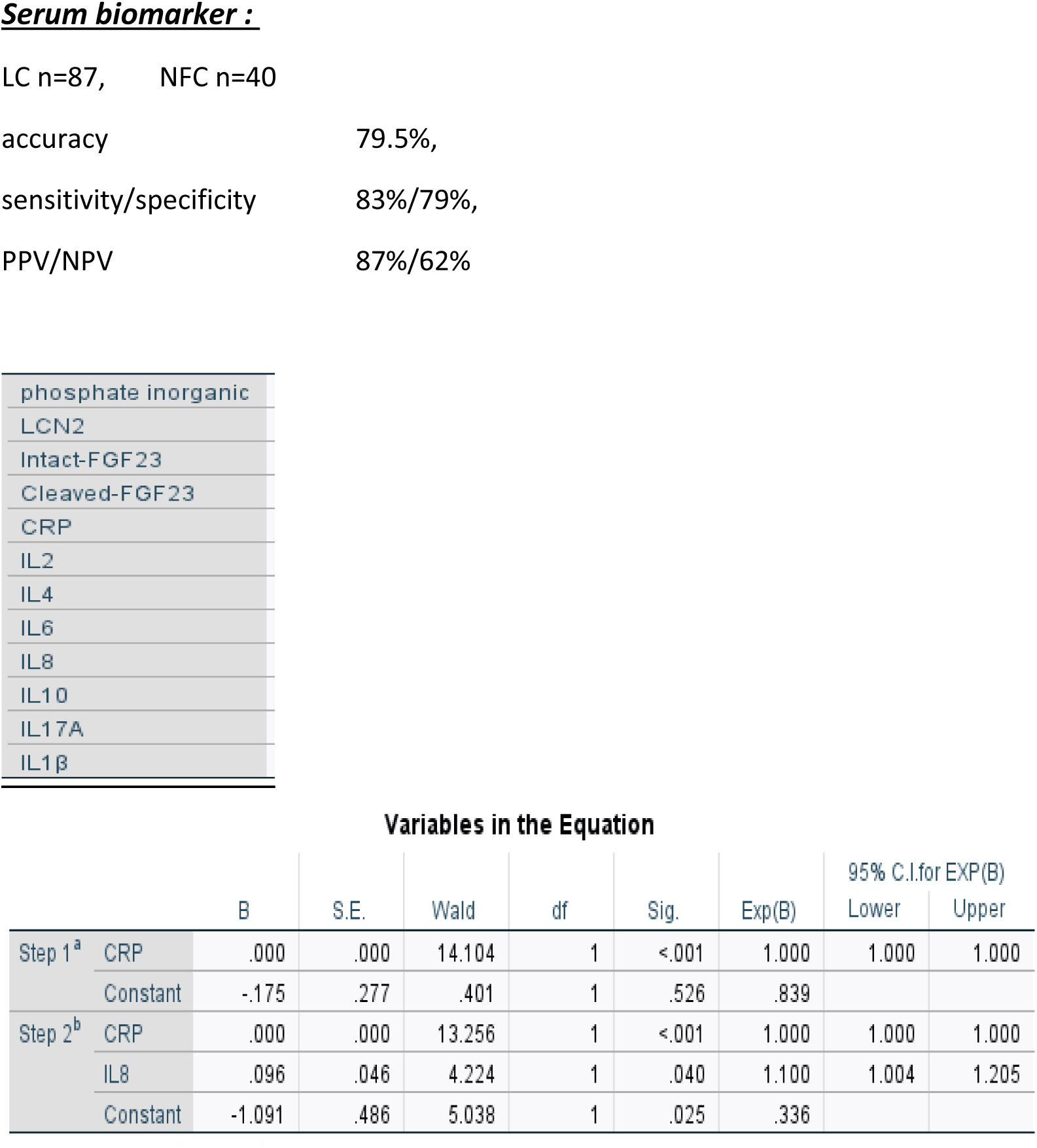
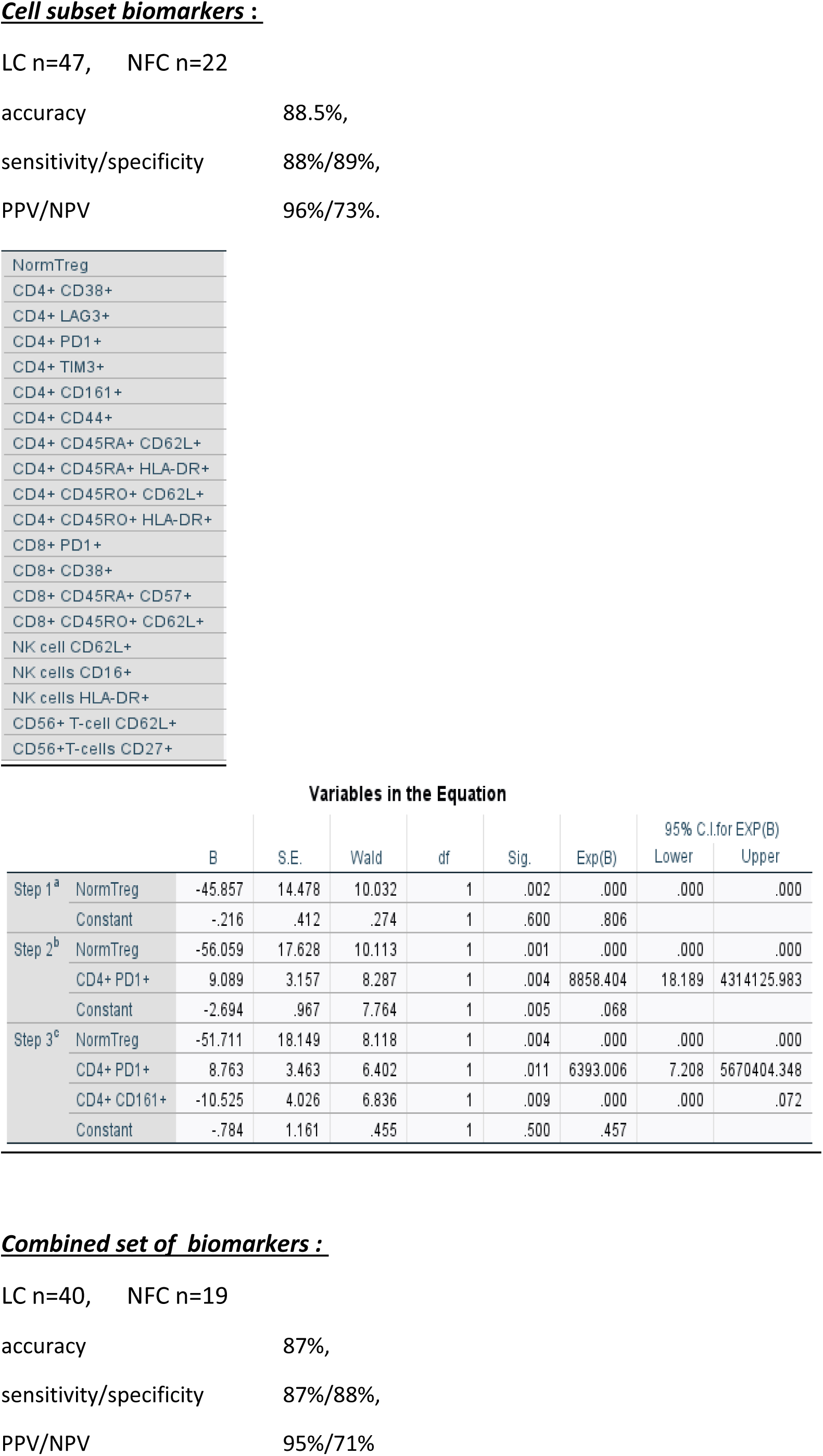
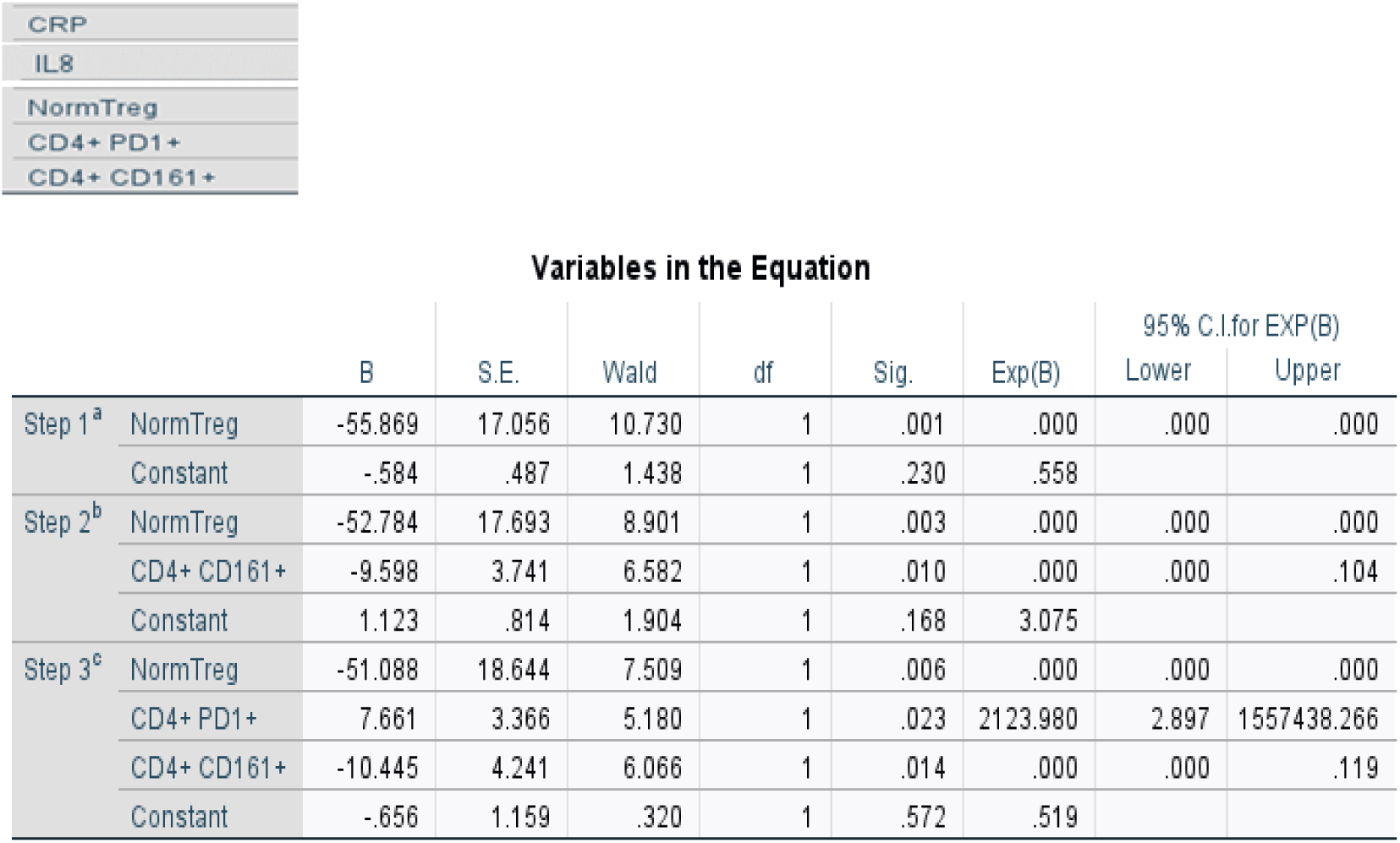
Regression analysis : LC versus NFC (Forward modelling approach)

**Figure-S11:**
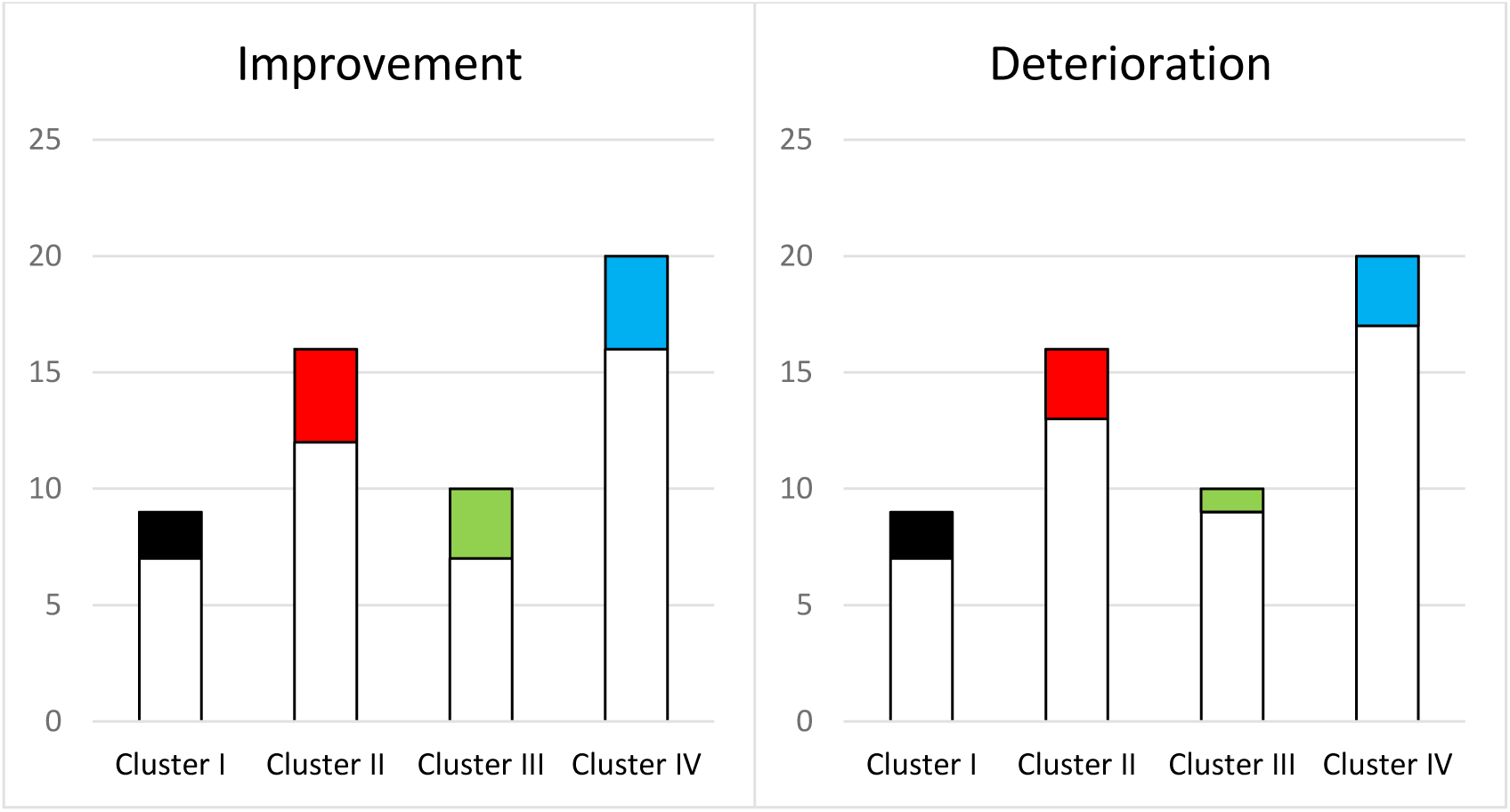
Cluster of patients defined in Figure-1 (n=55) were not associated with more patients showing improvement (n=13) or deterioration (filled histogram, n=13) over time compared to those with limited changes (open histograms).

##### Serum biomarker

##### Cell subset biomarkers

##### Combined set of biomarkers

